# Immunosenescence profile is associated with increased susceptibility to severe COVID-19

**DOI:** 10.1101/2024.11.18.24317502

**Authors:** Lucas Haniel A. Ventura, Lícia Torres, Giovanna Caliman Camatta, Jofer Zamame, Monique Macedo Coelho, Cecília Horta Ramalho-Pinto, João Gervazio, Felipe Caixeta, Leandro Nascimento, Mariana Almeida Oliveira, Vinícius Dantas Martins, Marcos Felipe Oliveira, Murilo Soares da Costa, Hugo Itaru Sato, Henrique Cerqueira Guimarães, Rafael Calvão Barbuto, Ana Paula Rocha Veiga, Najara Ataíde, Gabriela Prandi Caetano, Sarah Rangon, Mauro Lúcio O. Júnior, Fernanda Calvo Fortes, Luciana Zuccherato, Elaine Speziali, Olindo Assis Martins-Filho, Verônica Coelho, Roberto Avritchir, Rafael Souza, Marina Ayupe, Caio Loureiro, Maria Eduarda Passos, Ana Clara Mota Neves, Pauline Leite, Santuza Maria Ribeiro Teixeira, Unaí Tupinambas, Liza Figueiredo Felicori, Gabriela Silveira-Nunes, Tatiani Uceli Maioli, Denise Morais Fonseca, Andrea Teixeira-Carvalho, Ana Maria Caetano Faria

## Abstract

In this study we tested the hypothesis that the immunosenescence profile could account for the disproportional susceptibility of the elderly to severe forms of COVID-19. The immunological profiles of volunteers residing in endemic and non-endemic areas for chronic infectious diseases were analyzed at early stage of SARS-CoV-2 infection. A unique signature of inflammatory plasma mediators was identified in COVID-19 volunteers when compared to individuals with other flu-like syndromes. COVID-19 severity correlated with high levels of inflammatory mediators; among them, CXCL9, a serum marker of aging. Patients who progressed to hospitalization displayed high frequencies of CD8^+^ and CD4^+^ T cells expressing exhaustion and senescence markers and showed reduced and more mature B cell repertoires, which are typical of senescence. They also had an acceleration of epigenetic age measured by DNA methylation. Therefore, severe COVID-19 correlated with phenotypic, functional, and epigenetic features of accelerated immunosenescence at onset of infection.

## Introduction

Since the onset of COVID-19 cases, the elderly have been the main risk group for severe disease worldwide. This pattern of susceptibility was reported initially in China^1^, Italy^2^ and later in several countries including Brazil^3^.

SARS-CoV-2 infection is cleared by a normal anti-viral response in most individuals, but some progress to a hyperinflammatory condition, often with severe pulmonary involvement. It has been shown that 5–6% of these patients need mechanical ventilation due to respiratory failure developed by lung and microcirculation damage, with profound hypoxia, severe acute respiratory syndrome (SARS) and multiple organ dysfunction. These individuals have systemic hyperinflammation with classical serum biomarkers of “cytokine storm syndrome” including abnormal coagulation, elevated C reactive protein (CRP), LDH, ferritin, IL-1β, TNF, IL-6, IFN-γ, IL-10, IL-15, IL-2, IL-5, CCL2, CCL3, CCL4 and CXCL10 ^4^. Therefore, uncontrolled inflammation is clearly associated with severe life-threatening COVID-19. Interestingly, many of these mediators are the ones found to induce a low-grade chronic state of inflammation known as inflammaging and originated from senescent cells, cumulative stimulation of immune cells and lifestyle risk factors ^5,6^.

Aging affects all body compartments, but the immune system is particularly impacted. Immunosenescence is associated with a decline in the production and activity of T and B cells, many defects in innate immune cells, and thymic atrophy. The progressive reduction in the output of naïve T cells from the thymus creates an oligoclonal repertoire of expanded subsets of memory T cells and exhausted/senescent lymphocytes (mostly CD8 T cells) with low proliferative capacity. At the same time, senescent immune and non-immune cells develop a secretory phenotype (SASP) that includes innate cytokines and chemokines such as CXCL8, IL-6, IL-18, TNF and IL-1β responsible for the development of the inflammaging^6^. Immunosenescence is also associated with a reduction in production of B cells by the bone marrow. This decline in T/B cell production leads to the peripheral accumulation of cells with an exhausted repertoire displaying lower diversity of T/B cell receptors ^7,8^, and poor responses to infectious agents and vaccines ^6,9,10^. Thus, response to a virus infection such as SARS-CoV-2 in the elderly can be very abnormal as already observed for other viruses ^11^.

Interestingly, not all elderly developed the severe form of COVID-19 ^12^ and not all aged individuals have a dysfunctional immunological profile. Studies on healthy nonagenarians and centenarians in Italy and on healthy elderly people in Brazil showed that these individuals develop remodelling mechanisms to control the deleterious effects of inflammation and immunosenescence ^6,13–15^. On the other hand, our group have shown that individuals living in endemic areas for infectious diseases (such as Schistosomiasis, Leishmaniasis, Leprosy, Chagas Disease, Dengue, Chikungunya, Yellow Fever) exhibit an accelerated biological age as measured by DNA methylation^16^ with an unique immunological profile^14,17–20^. Therefore, aging does not affect individuals homogeneously and the resulting senescence phenotype seems to be determined by genetic, epigenetic and biographical factors related to the interactions of the immune system and the body with its environment.

Our hypothesis in this study is that the immunosenescence profile, rather than the chronological age, will impact on the development of the inflammatory damage seen in severe COVID-19 by creating a permissive environment for a dysfunctional response to the virus. Moreover, it is likely that factors such as chronic antigenic exposure in endemic areas for infectious diseases would enhance a preexisting immunosenescence phenotype.

Although some authors have theoretically proposed an association between COVID-19 severity and senescence biomarkers such as inflammaging, biological clocks, mTOR activity, and telomer shortening ^21–24^, only few studies have experimentally addressed this proposal using only cohorts of hospitalized individuals at heterogeneous time points of infection with mixed results ^25,26^.

Several distinctive parameters were used in this study to reach non-ambiguous conclusions when testing our hypothesis: (i) we investigated individuals from three cities in Brazil with distinct patterns of biological aging; (ii) our cohort had only individuals at early stage of infection (1-4 days of symptoms) to avoid confusion between the impact of immunosenescence in COVID-19 outcome with accelerated immunosenescence induced by SARS-CoV-2 infection itself ^27^; (iii) a group of individuals with flu-like symptoms but negative for SARS-CoV-2 infection was included as a control to distinguish COVID-19 from other airway viral infections; (iv) all individuals were clinically followed for 14 days to monitor disease progression, sometimes from mild to severe, as in the case of a specific group of patients

(progression group); (v) four parameters were tested to make up the immunosenescence profile - plasma mediators to examine the inflammaging, phenotypical biomarkers of memory, exhaustion and senescence in T cells, diversity and maturity of B cell repertoires and the epigenetic age of the individuals.

## Results

### Severe COVID-19 associated with a high inflammatory profile at early stage of infection

The main features of our study population are shown in **Table 1** and **2**. For all the analysis reported below, individuals in each group were matched by age, sex and number of comorbidities. We only included in the analysis individuals at early stage of infection (1-4 days of symptoms). Other factors that could interfere with the interpretation of our data were also controlled. (i) There was no significant difference in the body mass index (BMI) among the groups that could contribute as an inflammatory stimulus to the distinct outcomes ^28^. (ii) The presence of comorbidities was a general feature of our cohort, and the most reported ones were hypertension, diabetes mellitus, and respiratory diseases. Since they are inflammatory conditions that could interfere with COVID-19 severity ^29^, all groups analysed in this study were matched by the presence of an equal number of comorbidities (considered as equivalent conditions). (iii) To exclude the possibility that disease outcome would be related to differences in viral load as already reported for other infections by virus such as influenza ^30^ and adenovirus ^31^, concentration values for SARS-CoV-2 ribonucleic acid in each swab sample were calculate and compared among clinical groups. We used cycle threshold (Ct) values as a surrogate for viral load given the already reported linear correlation between Ct values and viral concentration down to the limit of detection^32^. **Supplementary Figure 1a** shows that individuals with mild, moderate, and severe COVID-19 presented comparable CT values indicating that disease outcome was not due to the abundance of virus in the respiratory tract. (iv) Individuals previously infected by SARS-CoV-2 were identified by serology (using Bio-Plex Multiplex SARS-CoV-2 Serology Assay Kit) and excluded from the study. (v) Since a high prevalence of Vitamin D deficiency has been reported to correlate with increased morbidity and mortality related to COVID-19 infection ^33^, serum levels of vitamin D were measured in individuals of our cohort. No significant difference among the clinical groups of COVID-19 was observed (**Supplementary Figure 1b**) ruling out the possible influence of its deficiency in the outcome of the disease.

**Table 1:** Characterization of the study sample.

|  | Flu-Like Syndrome<br>(n=131) | COVID-19<br>Mild (n=127) | COVID-19<br>Moderate (n=18) | COVID-19<br>Severe (n=33) | P value |
| --- | --- | --- | --- | --- | --- |
| **Age (years) (median; min/max)** | 38,0 (20±90) | 39,0 (20±77) | 49,0<br>(20±65) | **\*51,0 (31±68)** | **\*p=0,147** |
| **Sex** |  |  |  |  |  |
| Female (%) | 54,9 | 53,5 | 55,5 | 52,3 | - |
| Male (%) | 45,0 | 46,4 | 44,4 | 47,6 | - |
| **BMI (kg/m<sup>2</sup>) (median)** | 26,4 | 26,2 | 35,5 | 29,4 | p>0,05 |
| **Symptoms onset (days) (median)** | 4,0 | 4,0 | 5,0 | 5,0 | p>0,05 |
| **Comorbidities** |  |  |  |  |  |
| Hypertension (%) | 17,5 | 14,1 | 44,4 | 42,4 | - |
| Diabetes mellitus (%) | 0,7 | 3,1 | 16,6 | 15,1 | - |
| Respiratory diseases (%) | 10,6 | 11,0 | 0,0 | 6,0 | - |
| Others (%) | 10,6 | 12,5 | 5,5 | 18,1 | - |

Data presented as median (min-max) and mean ( ± standard deviation). The Kruskal-Wallis and Dunn's tests were applied. \*Corresponds to the difference found for comparison between the other compared groups.

**Table 2:** Characterization of the study sample by region.

| Belo Horizonte | Flu-Like Syndrome (n=76) | COVID-19 Mild (n=79) | COVID-19 Moderate (n=05) | COVID-19 Severe (n=14) | P value |
| --- | --- | --- | --- | --- | --- |
| **Age (years) (median; min/max)** | 41,0 (20±79) | 38,0 (20±68) | 51,0 (31±64) | **\*53,0 (32±89)** | **\*p=0,121** |
| **Sex** |  |  |  |  |  |
| Female (%) | 53,9 | 50,6 | 20,0 | 28,5 | - |
| Male (%) | 46,0 | 49,3 | 80,0 | 71,4 | - |
| **BMI (kg/m2) (median)** | 26,1 | 26,0 | 33,5 | 28,4 | p>0,05 |
| **Symptoms onset (days) (median)** | 4,0 | 5,0 | 2,0 | 4,0 | p>0,05 |
| **Comorbidities** |  |  |  |  |  |
| Hypertension (%) | 11,8 | 11,3 | 20,0 | 14,2 | - |
| Diabetes mellitus (%) | 2,6 | 3,7 | 20,0 | 14,2 | - |
| Respiratory diseases (%) | 10,5 | 11,3 | 0,0 | 0,0 | - |
| Others (%) | 7,8 | 13,9 | 0,0 | 7,1 | - |
| Governador Valadares | Flu-Like Syndrome (n=44) | COVID-19 Mild (n=42) | COVID-19 Moderate (n=00) | COVID-19 Severe (n=03) | P value |
| **Age (years) (median; min/max)** | 35,0 (20±90) | 39,0 | - | 45,0 | p>0,05 |
| **Sex** |  |  |  |  |  |
| Female (%) | 47,7 | 59,5 | - | 33,5 | - |
| Male (%) | 52,3 | 40,4 | - | 66,5 | - |
| **BMI (kg/m2) (median)** | 26,6 | 26,2 | - | 22,3 | p>0,05 |
| **Symptoms onset (days) (median)** | 3,0 | 4,0 | - | 3,0 | p>0,05 |
| **Comorbidities** |  |  |  |  |  |
| Hypertension (%) | 9,0 | 11,9 | - | 33,3 | - |
| Diabetes mellitus (%) | 0,0 | 0,0 | - | 0,0 | - |
| Respiratory diseases (%) | 4,54 | 4,7 | - | 0,0 | - |
| Others (%) | 0,0 | 2,3 | - | 33,3 | - |
| São Paulo | Flu-Like Syndrome (n=11) | Covid-19 Mild (n=06) | Covid-19 Moderate (n=13) | Covid-19 Severe (n=16) | p value |
| **Age (years) (median; min/max)** | 40,0 (23±69) | 41,5 (28±50) | 48,0 (20±56) | 51,0 (31±64) | p>0,05 |
| **Sex** |  |  |  |  |  |
| Female (%) | 45,4 | 50,0 | 69,2 | 56,2 | - |
| Male (%) | 54,5 | 50,0 | 30,7 | 43,7 | - |
| **BMI (kg/m2) (median)** | - | - | - | - | - |
| **Symptoms onset (days) (median)** | 5,0 | 4,5 | 5,0 | 5,0 | p>0,05 |
| **Comorbidities** |  |  |  |  |  |
| Hypertension (%) | 9,9 | 0,0 | 38,4 | 37,5 | - |
| Diabetes mellitus (%) | 0,0 | 0,0 | 7,6 | 18,7 | - |
| Respiratory diseases (%) | 27,7 | 0,0 | 0,0 | 12,5 | - |
| Others (%) | 0,0 | 0,0 | 7,6 | 12,5 | - |

Data presented as median (min-max) and mean ( ± standard deviation). The Kruskal-Wallis and Dunn's tests were applied. \*Corresponds to the difference found for comparison between the other compared groups.

Since previous studies have shown that inflammaging is a hallmark of immunosenescence^10^, we first characterized the inflammatory profile of the individuals in all groups using a Luminex-Multiplex assay for 27 immune mediators. For this first analysis, the study population was segregated into five groups: Flu-like Syndrome (FLS) group comprising individuals who were negative for SARS-CoV-2 and had an unknown mild respiratory infection, Mild, Moderate and Severe COVID-19 groups according to WHO classification guidelines^34^ (**Supplementary Table 1**). The fifth group (Progression) consisted of individuals who were classified as Mild COVID-19 at recruitment but progressed to severe disease with hospitalization during the 14-day follow-up period.

A radar chart showing a global analysis of plasma mediators produced by individuals with mild, moderate, and severe COVID-19 shows that, at early stage of infection, disease severity was already positively correlated with the frequency of high producers of several inflammatory mediators (**Figure 1a**). Among these mediators, we identified 10 that were significantly different among the groups (**Figure 1b**): CCL2, CXCL8, CXCL10, IL-1β, IL-6, TNF, IFN-gamma, IL-12p70, IL-1Ra, IL-10. These are classical inflammatory mediators that are part of the inflammaging phenotype ^10,35^ and were already reported to be at high levels in hospitalized COVID-19 patients. Interestingly, the same list of mediators was also significantly higher in individuals with mild COVID-19 when compared to the ones with flu-like syndrome (**Figure 1b**). Although individuals from these two groups shared a spectrum of symptoms and could only be segregated after RT-PCR testing for SARS-CoV-2 infection, they had a distinct inflammatory profile. Mild COVID-19 patients presented a higher global pattern of mediator production (**Figure 1c**) and several differences in the concentrations of the 10 mediators (**Figure 1d, Supplementary Figure 2c**) since early infection. We also observed differences in this inflammatory profile at other time points of disease progression with higher immune activation in mild COVID-19 patients when compared to FLS patients with 5-9 and 10-14 days of symptoms (**Supplementary Figure 2 d, e**). Probably at this stage, immune mediators resulting from the response to infection are at play. Therefore, COVID-19 seems to be related to a unique inflammatory profile when compared to other respiratory viral infections with similar symptoms. Notably, when patients with mild COVID-19 were segregated into adults and elderly, a higher global profile of inflammatory immune activation was detected in aged individuals (**Supplementary Figure 2b, e**) during early stages of infection (1-4 days of symptoms) and later (10-14 days of symptoms) suggesting that inflammaging and the innate immune response triggered by COVID-19 indeed overlap later when immunity to infection is stablished.

**Figure 1:**
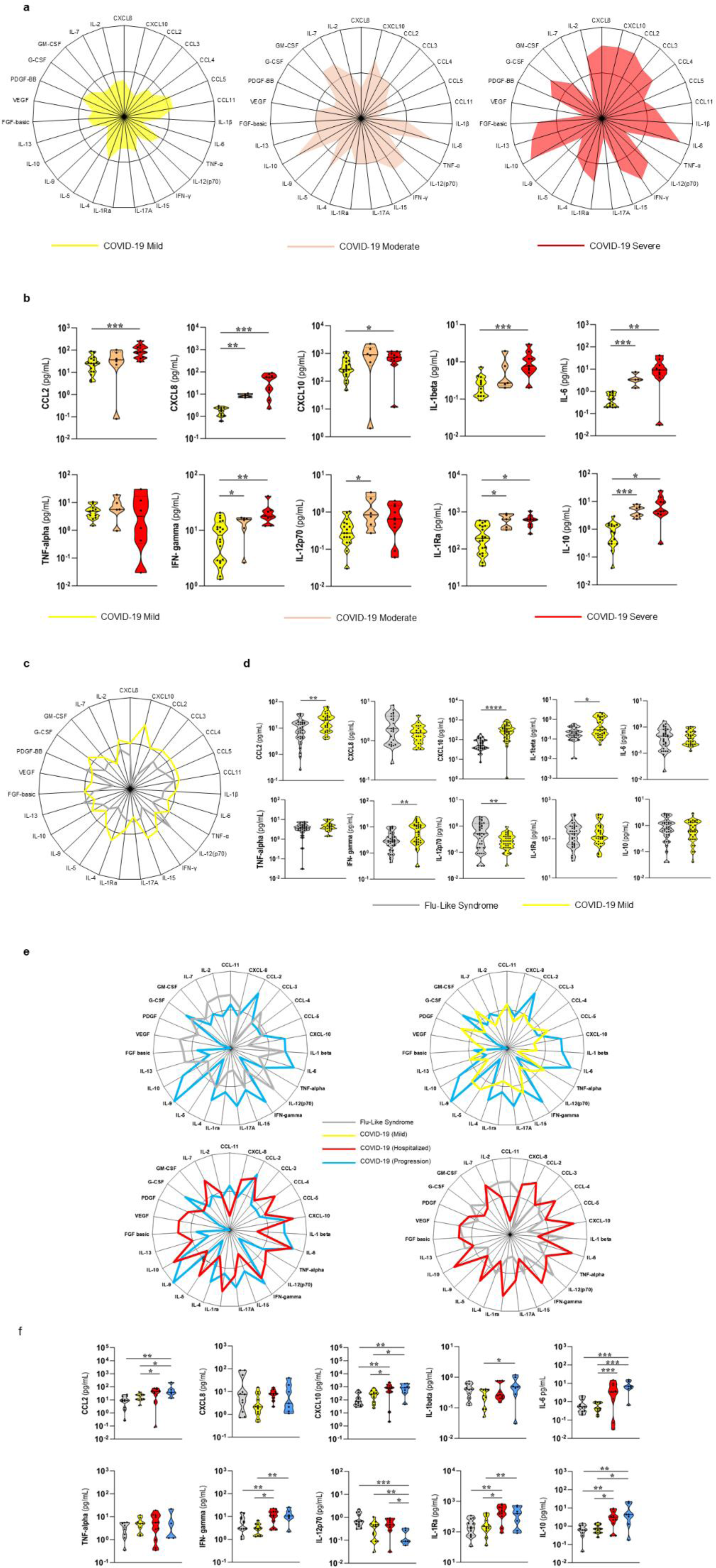
Early inflammatory profile of COVID-19 and Flu-Like syndrome patients. **(a)** Radar chart with the frequency of high mediator producers among individuals infected with COVID-19 from Belo Horizonte and São Paulo (n=36), Mild COVID-19 (n=20), Moderate COVID-19 (n=6) and Severe COVID-19 (n=10). **(b)** Differences in plasma mediator concentrations in individuals with different clinical forms of COVID-19. The population used for this analysis was the same as that used in Figure 1A. Samples were previously normalized, and outliers were excluded using the ROUT test. Mann-Whitney test was performed individually for each group. Lines were used to represent which groups were compared and asterisks to represent statistical significance (p≤0.05, **p≤0.01, ***p≤0.001 or ****p<0.0001). **(c)** Radar chart representing the frequency of high mediator producers among individuals infected with COVID-19, using individuals from Belo Horizonte and São Paulo (n=74), Flu-Like Syndrome (n=38), Mild COVID-19 (n=42). **(d)** Comparative analysis of the concentration of each mediator among groups. Samples were previously normalized, and outliers were excluded from the data, using the ROUT test. Mann-Whitney test was performed individually for each group. Asterisks to represent statistical significance (p≤0.05, **p≤0.01, ***p≤0.001 or ****p<0.0001). **(e)** Radar charts representing the frequency of high producers of plasma mediators in individuals from Belo Horizonte (n=46) with Flu-Like Syndrome (n=13), Mild COVID-19 (n=14), Hospitalized COVID-19 (n=12) and COVID-19 Progression (n=7). The Hospitalized COVID-19 group consists of individuals who were hospitalized with either moderate or severe COVID-19. **(f)** Analysis of the concentration of plasma mediators of individuals from Belo Horizonte with either COVID-19 or Flu-Like Syndrome (FS). The population used for this analysis was the same as that used in D. Asterisks represent statistical significance (*p≤0.05, **p0.001, ***p0.001 or ****p0.0001). All groups presented in this figure were matched by sex as well as age and they were composed of individuals at the initial stage of infection (1-4 days of symptoms).

To help distinguishing between pre-existing inflammaging as a fuel to disease progression and the viral-induced cytokine storm, we studied an important group of patients found in our cohort: the progression group. These individuals were classified as having mild symptoms of COVID-19 when they were recruited (1-4 days of symptoms) but progressed to severe disease and hospitalization during the 14-day-follow-up period. Notably, the overall inflammatory profile of these patients represented by a radar plot (**Figure 1e**) and the comparative analysis of the 10 main plasma mediators (**Figure 1f**) clearly showed that they presented, since the early stages of infection, an inflammatory activation similar to the group of severe COVID-19 patients, and significantly different from Mild and FLS patients. This suggests that the initial immunological imprint of inflammatory mediators can predict disease outcome.

### Severe COVID-19 was associated with higher inflammatory activation in individuals from endemic areas for infectious diseases and with biomarkers of aging

To address the question of how distinct environments influence COVID-19 and whether aging biomarkers can be used to distinguish the different clinical forms of disease, we compared the inflammatory profile of adult and elderly individuals with FS or mild COVID-19 from Belo Horizonte and Governador Valadares, an endemic area for several infectious diseases where individuals present an accelerated biological aging ^16^. First, we confirmed that there was a clear difference in the global inflammatory profiles between individuals with FLS and Mild COVID-19 in the endemic area despite their similarity of symptoms (**Figure 2c**). Moreover, individuals from Governador Valadares had a higher activation profile when compared to individuals from Belo Horizonte, and the elderly from the endemic area presented a much higher production of immune mediators than adults (**Figure 2c**). Hence, not only does COVID-19 presented itself as a greater challenge to the immune system than other respiratory infections, but the inflammatory activation at early phase of infection is exacerbated in populations with accelerated biological age.

**Figure 2:**
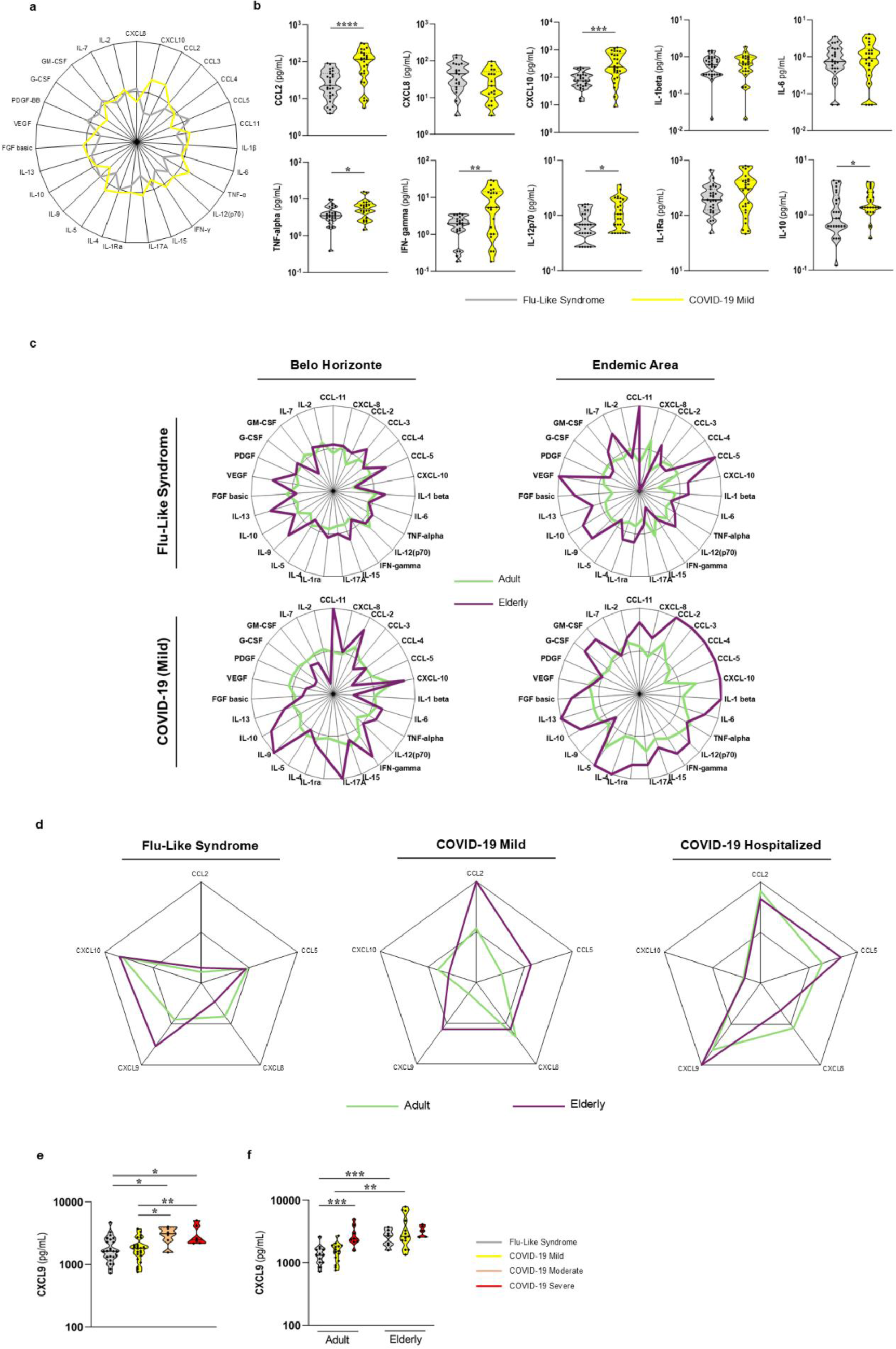
Inflammatory profiles of adult and elderly people with Flu-like syndrome and COVID-19 living in endemic and in non-endemic areas for infectious diseases. **(a)** Radar chart representing the frequency of high producers of mediators among individuals from Governador Valadares (n=56) with either Flu-Like Syndrome (n=30) or Mild COVID-19 (n=26). **(b)** Comparative analysis of concentrations of plasma mediators in individuals with either Flu-like syndrome or Mild COVID-19. Samples were previously normalized, and outliers were excluded using the ROUT test. Mann-Whitney test was performed individually for each group. Asterisks represent statistical significance (p≤0.05, **p≤0.01, ***p≤0.001 or ****p<0.0001). **(c)** Radar charts representing the frequency of high producers of mediators among individuals with COVID-19 in the adult (20-59 years) and elderly (over 60 years) age groups in a non-endemic area (Belo Horizonte and São Paulo) - Adults (n=92) with Flu-Like Syndrome (n=45) and Mild COVID-19 (n=47) and elderly (n=19) with Flu-Like Syndrome (n=11) or Mild COVID-19 (n=8) and in an endemic area (Governador Valadares) - Adults (n=75) with Flu-Like Syndrome (n=39) and Mild COVID-19 (n=36) and elderly (n=11) with Flu-Like Syndrome (n=5) and Mild COVID-19 (n=6) **(d)** Radar charts representing the frequency of high producers of CXCL9 among all individuals with Flu-Like Syndrome and COVID-19 in the adult and elderly age groups - (Adults n=40) with FS (n=14), Mild COVID-19 (n=15) and Hospitalized (n=11), elderly (n=26) with FLS (n=13), Mild COVID-19 (n=7) and Hospitalized (n=6). **(e)** Comparative analysis of plasma concentrations of CXCL-9 in individuals with different clinical forms of COVID-19. Samples were previously normalized, and outliers were excluded using the ROUT test. Mann-Whitney test was performed individually for each group. Asterisks represent statistical significance (p≤0.05, **p≤0.01, ***p≤0.001 or ****p<0.0001). **(f)** Comparative analysis of plasma concentrations of CXCL-9 in individuals with different clinical forms of COVID-19. The population used is the same shown in Figure 2C. Samples were previously normalized, and outliers were excluded from the data using the ROUT test. Thus, the Mann-Whitney test was performed individually for each group. Lines were used to represent which groups were compared and asterisks to represent statistical significance (p≤0.05, **p≤0.01, ***p≤0.001 or ****p<0.0001). All groups presented in this figure were matched by sex as well as age and they were composed of individuals at the initial stage of infection (1-4 days of symptoms).

CXCL9 has been recently identified as critical mediator involved in age-related chronic inflammation ^36^. Indeed, using this chemokine as a biomarker of aging and frailty, we observed that elderly individuals were higher producers of CXCL9 than adults. Interestingly, both adults and elderly hospitalized patients had a similar and more prominent production of this mediator than individuals with FLS or mild COVID-19 (**Figure 2d**). Moreover, elderly individuals presented no difference in CXCL9 production across the spectrum of COVID-19 clinical forms (**Figure 2e**) probably because they already had high baseline levels of this chemokine. Therefore, severe COVID-19 was associated at early stage of infection with increased production of an immunological biomarker of aging.

Our data on the inflammatory profile of individuals from endemic and non-endemic areas in Brazil suggest a strong and unique association between inflammaging at early stages of COVID-19 to disease progression.

### Severe COVID-19 correlated with accumulation of memory, exhausted and senescent T cells

To further explore the association between other features of immunosenescence and COVID-19 outcomes, we investigated changes in the frequencies of T cell populations expressing cell markers of memory, exhaustion, and senescence.

Aging has a great impact in the T cell compartment, especially in CD8^+^ T cells^37^. To investigate variations in the frequencies of functional subsets of T lymphocytes, multiparametric flow cytometry analyses was performed using a panel of monoclonal antibodies to surface markers of exhaustion/senescence in these cells (**Supplementary Table 2**). Gating strategies for all cell populations are shown in **Supplementary Figure 3**.

To investigate immunophenotypes of T lymphocytes, CD4^+^ and CD8^+^ T cells were studied as global main subpopulations and segregated into functional subsets of naïve (CD45RO^-^CCR7^+^), effector (CD45RO^-^CCR7^-^), central memory (CD45RO^+^CCR7^+^), effector memory (CD45RO^+^CCR7^-^) T cells. We identify accumulation of several subsets of CD8^+^ and CD4^+^ T cells expressing markers of exhaustion and senescence in patients with severe COVID-19 (**Figure 3**). Concerning CD8^+^ T cell analysis, individuals with severe disease had higher frequencies of CD8^+^ T cells expressing biomarkers of senescence (TIGIT) and exhaustion (ICOS). Effector CD8^+^ T cells expressing PD-1 were also augmented in severe diseased patients (**Figure 3 a, b**). In addition, CD8^+^ T cells with senescence/ exhaustion phenotypes accumulate in individuals of this group: CD28-PD-1^+^CD57^+^CD8^+^, CD28^+^KLRG1^+^CD8^+^, CD28^-^PD-1^+^CD8^+^, CD28^-^KLRG1^+^ PD-1^+^ CD8^+^, KLRG1^+^PD-1^+^CD8^+^, KLRG1^+^PD-1^+^CD8^+^, TIGIT^+^ICOS^+^ CD8^+^ and CD28-PD-1^+^ effector and effector memory CD8^+^ T cells (**Figure 3 c, d**). For CD4^+^ T cells, we also found higher frequencies of cells expressing the senescence marker TIGIT and of cells expressing the mixed senescence/exhaustion phenotype: TIGIT^+^ICOS^+^CD4^+^, CD28^-^CD57^+^KLRG1^+^CD4^+^ and CD28^-^CD57^+^KLRG1^+^ effector memory CD4^+^ T cells (**Figure 3e, f**). For some specific sub-populations of senescent (CD4^+^TIGIT^+^) (**Figure 3e**) and senescent/exhausted (CD28^-^PD-1^+^ effector and effector memory CD8^+^, TIGIT^+^ICOS^+^CD4^+^) T cells (**Figure 3c**), a significant difference between patients with mild and moderate disease was observed. Therefore, along with an exuberant inflammaging, COVID-19 patients exhibited an accrual of senescent/exhausted CD8^+^ and CD4^+^ T cells that make up the scenario of dysfunctional immunity permissive to disease progression.

**Figure 3:**
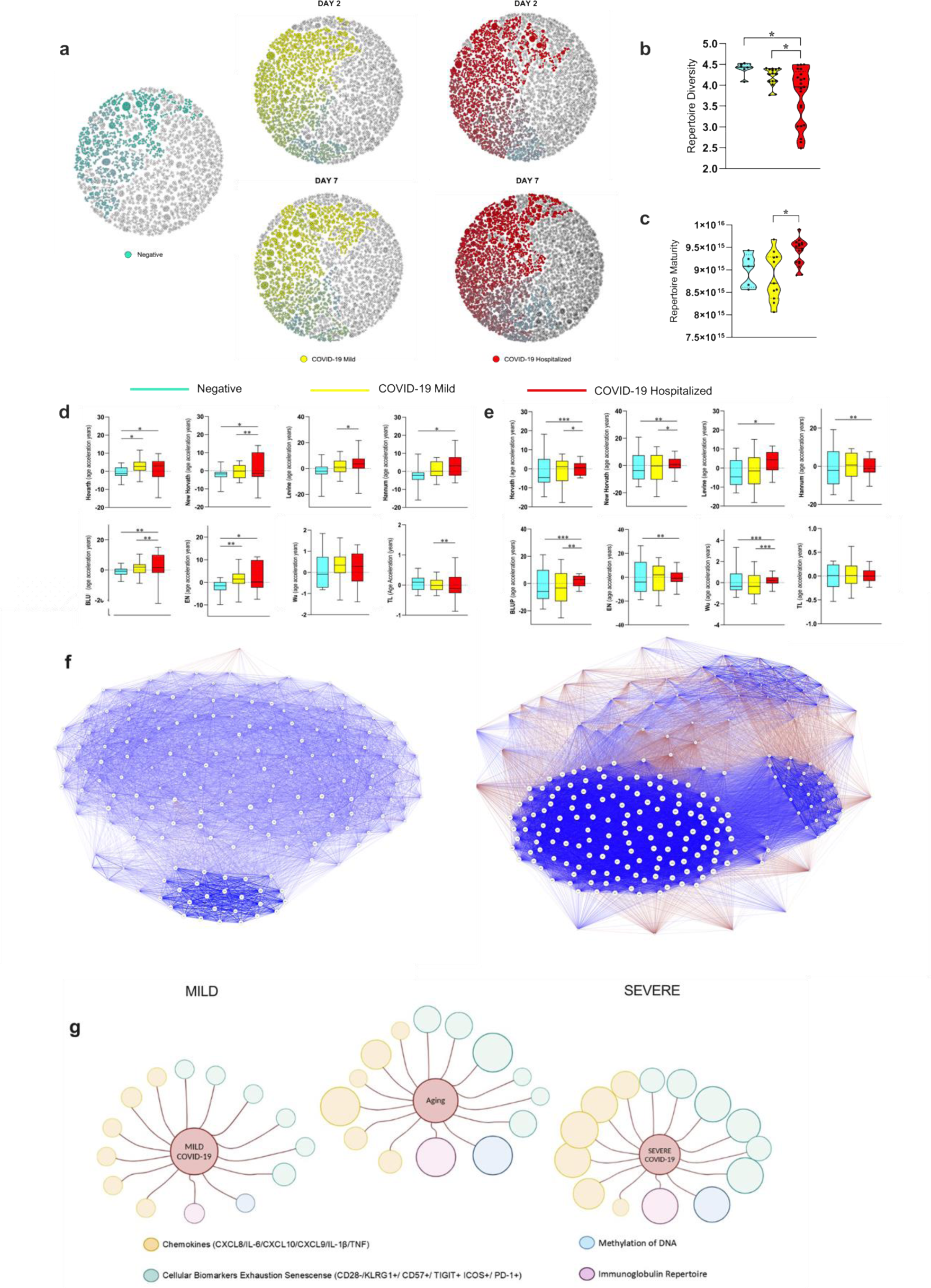
The senescence and exhaustion profile was significantly increased in individuals with moderate or severe COVID-19 compared to mild individuals in both CD8^+^ and CD4^+^ T cells. Patients were also stratified into 3 groups classified according to the severity of the disease as mild (n=20), moderate (n=8) and severe (n=8). (**a**) Percentage of senescent and exhausted CD8^+^ T cells from individuals with mild, moderate and severe COVID. (**b**) The gating strategy was used to analyze markers related to senescence and exhaustion together within CD8^+^ T cells. Senescence cells are TIGIT^+^; exhausted are PD-1^+^ and ICOS^+^. (**c**) Percentage of senescent/exhausted CD8^+^ T cells from individuals with mild (20), moderate (8) and severe (8) COVID-19. Senescent/exhausted CD8^+^ T cells quantified by co-expression of CD57^+^ and PD-1^+^, CD28neg and PD-1^+^, TIGIT^+^ and ICOS^+^. KLRG1^+^ marker may be present in senescent and exhausted. (**d**) Representative flow cytometry of senescent/exhausted CD8^+^ T cells, with the gate of interest showing a red square. (**e**) Percentage of senescent and exhausted CD4^+^ T cells from individuals with mild, moderate and severe COVID. (**f**) Dot plots of strategy was used to analyze markers related to senescence TIGIT^+^, CD57^+^, CD28neg and KLRG1^+^, to exhaustion marker ICOS^+^ and KLRG1^+^ within CD4^+^ T cells was used (red square). *p < 0.05, **p < 0.01.

### Severity of COVID-19 was associated with a reduction in B cell diversity and higher maturation of the immunoglobulin repertoire

To study the B cell senescence, we examined the immunoglobulin repertoire in a set of 45 individuals from individuals in Belo Horizonte and Governador Valadares (endemic area) who were either uninfected or tested positive for COVID-19. Within these cities, participants were categorized into negative group (n=6), mild COVID-19 group (n=19), and hospitalized COVID-19 group (n=20) of individuals from Belo Horizonte and São Paulo along with mild COVID-19 individuals from Governador Valadares. The sequencing resulted in 285,341 to 1,525,890 raw reads. After pre-processing, 130,060 to 548,060 annotated reads and 3,053 to 69,737 clones were obtained (**Supplementary Table 3**).

Antibody repertoire diversity within the top 100 most expanded clones among individuals in Belo Horizonte showed that the Shannon entropy diversity was lower in the hospitalized individuals as compared to both mild COVID-19 and control groups (**Figure 4b**). **Figure 4c** also shows that hospitalized individuals had a more mature repertoire with highly mutated VH immunoglobulin genes and lower frequencies of germline-encoded antibodies, a phenotype of an aged repertoire. A graphical representation of clonal expansion at day 2 and 7 of symptom onset shows that in both periods the clones from severe individuals are distinct from baseline (negative) and clones from mild individuals (**Figure 4a**).

**Figure 4:**
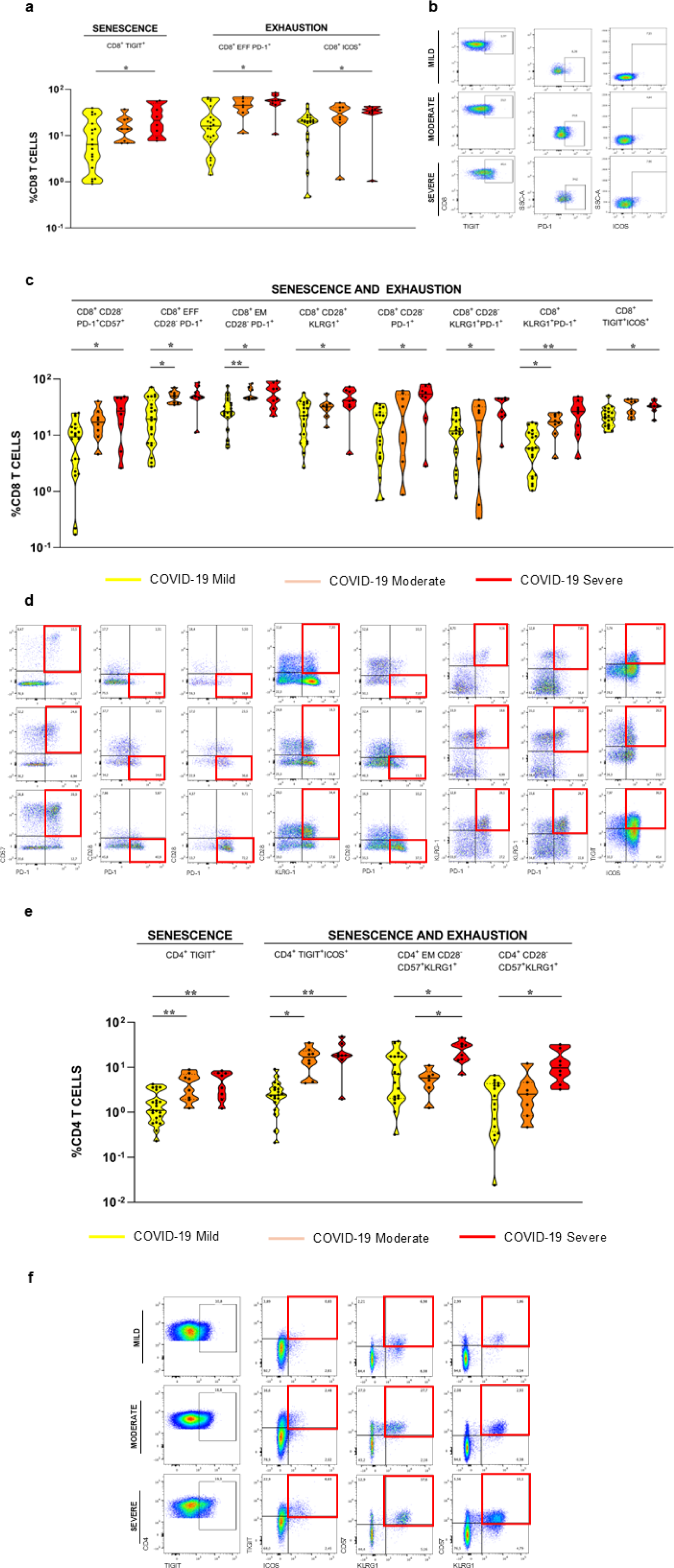
Analysis of immunoglobulin repertoires and of biological aging in patients with different clinical forms of COVID-19. **(a)** Graphical representation of clonal expansion of the 100 most expanded clones in the repertoire of five individuals. The more intense the color, the more expanded the clone is. **(b)** Shannon entropy diversity was calculated using the diversity function of the ‘vegan’package with the 100 most expanded clones for hospitalized individuals (n=20, Red), individuals with mild cases (N=19, Yellow), and negative individuals (n=6, Blue-Cyan) from Belo Horizonte and Governador Valadares (only mild cases). Differences among groups were calculated using Mann-Whitney test and ROUT test to exclude outliers. Asteriscs represent statistical significance (*p<0.01). (**c**) Repertoire maturity (divergence from germline-like sequences) was calculated for the whole repertoire of hospitalized individuals (n=20, Red), individuals with mild cases (N=19, Yellow), and negative individuals (n=6, Blue-Cyan) from Belo Horizonte and Governador Valadares (only mild cases). The ROUTE test was performed to remove outliers. **(d)** Accelerated aging was measured using eight different epigenetic clocks (Horvath; New Horvath; Levine; Hannum; BLUP; EN; Wu; TL) in unvaccinated COVID-19 negative control individuals (n = 12) (Blue-Cyan), infected with COVID-19 Mild 9 (n = 9) (Yellow) and infected with Hospitalized COVID-19 (n = 17) (Red) without including endemic area. The y-axis shows the acceleration of epigenetic age after adjustment for cell type fractions (Residual Regression of Epigenetic Age Acceleration on Cell Type Fractions). The box plot represents the median, interquartile, minimum and maximum values of age acceleration (in years) determined for each group. One-way ANOVA and t-test were used to evaluate significant differences. Values of p < 0.05 were considered statistically significant (*). **(e)** The same analysis as in Figure 4D was performed but including individuals from endemic areas. The population used includes unvaccinated COVID-19 negative control individuals (n = 12) (Blue-Cyan), infected with COVID-19 Mild 9 (n = 18) (Yellow) and infected with COVID-19 Hospitalized (n = 18) (Red). **(f)** Biological network of cytokines and cellular proteins of exhaustion and senescence in infected individuals with mild and severe COVID-19. Patients who developed severe COVID-19 showed greater interactions with biomarkers of senescence and exhaustion in immune cells (including cytokines, chemokines, and cell surface markers) up to 4 days after infection, when compared to patients who developed the mild form of the disease. Blue thicker lines indicate positive correlations and red thicker lines negative correlations. The description of the codes present in the graph is in Supplementary Table 5. The size of the circles is proportional to their importance as hubs of connection. **(g)** A graphical representation of the main biomarkers of aging (cytokines/chemokines, Ig repertoire, DNA methylation, cellular markers of exhaustion and senescence) illustrates the comparative differences between individuals with Mild and Severe COVID-19 comparing them with the same markers described for aged individuals. The size of the circles is proportional to the intensity of the parameter they represent.

### Acceleration of biological age was detected in hospitalized individuals with moderate/severe COVID-19

To further explore our hypothesis, we accessed the DNA methylation status of peripheral blood mononuclear cells to investigate whether COVID-19 severity was associated with acceleration of biological age. We used the cohort of individuals that had been already investigated for the inflammatory and phenotypic lymphoid profiles and performed the analysis using different biological age acceleration clocks ^16,38^.

For this analysis, a negative population was selected as a control. Individuals in this group had no symptoms of infection and were negative for SARS-CoV-2 by RT-PCR (**Table 1**). Negative individuals were compared to infected patients with Mild COVID-19 and a Hospitalized COVID-19 group of patients with either moderate or severe clinical disease. Several clocks are now available to access aging acceleration using different gene clusters. We used seven clocks proposed by Horvath ^38^, Horvarth Updated, Levine ^39^, Hannum ^40^, TL ^41^, BLUP, EN, Wu ^25,42^, to obtain more accuracy in detecting acceleration or deceleration of biological age.

Based on data previously published by our group showing that the population of Governador Valadares presents an acceleration of biological age ^16^, we decided not to use these individuals in the initial analysis, so that we could focus on understanding the association between age acceleration and disease severity (**Figure 4D**). Notably, individuals of the hospitalized group had an increase in biological age when compared to the mild COVID-19 group (in Horvarth (Updated), Levine, EN, BLUP and TL watches) and to the negative group (in Horvarth, Horvarth (Updated), Hannum, BLUP and EN watches) (**Figure 4D**). The Wu clock was the only one of the analysed watches that showed no difference among groups. Next, we analyzed the total population, including individuals from Governador Valadares, and the result was similar (**Figure 4E**) except for the TL clock that showed no difference among groups.

Network analysis is a powerful tool for understanding the complex relationships between variables within a system. By constructing a network representation, we can visualize and analyze the interconnections between different components. In our study, we employed a weighted network analysis to investigate the correlations between biologically relevant markers associated with cytokines/chemokines and cellular proteins that are markers of senescence and exhaustion. Individuals who developed severe COVID-10 had a more connected senescence/exhaustion network than individuals that developed mild COVID-19 (**Figure 4F**). Among the more connected clusters, senescent and exhausted T cells are dominant. A graphical representation of all data obtained on the senescence profile illustrates the general conclusion that patients with severe disease resemble the aging phenotype (**Figure 4G**).

Therefore, our data clearly correlate several features of the immunosenescence profile including inflammaging, increased frequencies of senescent/exhausted T cells, oligoclonality of B cell repertoire and finally an acceleration of biological age suggesting that this dysfunctional immune profile can explain, at least in part, the high susceptibility of aged people to develop severe forms of COVID-19.

## Discussion

Since the beginning of the SARS-CoV-2 pandemic, the elderly have been the main risk group for the mortality and the severe clinical outcomes of COVID-19 ^2^. Several review articles have proposed that remodelling features acquired by the aging immune system such as the inflammaging phenomenon and dysfunctional immunity might have a major role in the unfavourable outcome of the disease in older individuals ^4,21–24,43^. Besides these theoretical proposals, some studies have addressed the hypothesis in experiments using senolytics in aged mice infected with a SARS-CoV-2– related virus^44^ and in individuals with severe and critically ill COVID-19 patients by analyzing exhausted/senescent T cells ^45,46^ and age-acceleration clocks ^25^.

In our study, we used more than one approach to reach a reliable conclusion when testing our hypothesis, we distinguish patients with mild disease from those severely ill to investigate the effect of aging phenotype in the disease outcome and we specifically restrict our sample to early infected patients (1-4 days of symptoms), a period when senescent and exhausted T cells are at baseline frequencies (a small percentage of the overall cell population) in mild COVID-19^27^. Our group and others have shown that SARS-CoV-2 infection induces not only a systemic inflammatory response similar to the inflammaging^47^ but it also leads to the emergence of exhausted and senescent T cells after a period of infection as short as 7 days ^27,45^. In addition, we included volunteers with mild flu-like symptoms who tested negative for SARS-CoV-2 as a control since these individuals did not show accumulation of exhausted/senescent cells even after 7 days of infection. At this stage, there is probably an overlap between inflammaging and innate cytokines/ chemokines produced in the early response to SARS-CoV-2 ^47^ because these two responses share a set of mediators including IL-6, CXCL8, C-Reactive Protein (CRP), TNF, IL-1β, IL-10, IL-1Ra IL-12p70, IFN-γ ^6,10^.

Ten mediators were consistently identified in all COVID-19 patients with 1-4 days of symptoms, and they were detected at variable levels depending on the clinical form of the disease. Most of them are proinflammatory mediators (CCL2, CXCL8, CXCL10, IL-1β, IL-6, TNF, IFN-γ, IL-12p70) that contribute to both frailty in the elderly ^10,36,48^ and to a series of pathological events seen in COVID-19 such as thrombosis of blood capillaries, congestion of the alveolar wall, inflammatory infiltration, formation of hyaline membrane in the alveoli, pulmonary edema, and severe respiratory failure ^5,49^ whereas others play an anti-inflammatory and regulatory role (IL-1Ra and IL-10) in both conditions. Interestingly, even though other viral infections can induce a burst of innate cytokines/chemokines during the first few days of infection ^47^, the inflammatory signature observed in individuals with mild COVID-19 and Flu-like syndrome in our study were not equivalent (**Figure 1c, d, Supplementary Figure 1**). This finding underscores the uniqueness of SARS-CoV-2 infection and suggests that the overlap between innate response to the virus and the pre-existing inflammaging phenomenon is of particular importance to COVID-19 with predictive value for the outcome of the disease. A further demonstration of that is the data collected from individuals in the Progression group (**Figure 1e, f**) that clearly showed an inflammatory signature of severe disease at early stages of infection when symptoms are still mild.

Another distinctive feature of our study population was the inclusion of a group of patients from an endemic region for infectious diseases in Brazil (Governador Valadares, MG). This group allowed us to examine the impact of accelerated imunosenescence and other changes favourable to the senescence process ^16,18^ in COVID-19 development. Indeed, we found a more prominent inflammatory profile in patients from the endemic area even in the mild COVID-19 and flu-like syndrome groups and this hyperactivation was exacerbated in the elderly (**Figure 2 a-c, Supplementary Figure 1**). These set of results highlight two important aspects concerning infectious diseases. First, it demonstrated that there is a variability of immune responses to a single pathogen, even for those with the same spectrum of symptoms, and this may be determined by genetic and environmental conditions as previously proposed ^50^. In addition, it suggests that individuals living in areas of high antigenic burden, despite their accelerated senescence profile ^16^, may also develop mechanisms of inflammatory remodelling resembling the ones observed in centenarian populations of Italy ^6,48^ and aged populations in Brazil ^17,37^. These mechanisms would allow them to successfully cope with life-threatening infectious disease such as COVID-19.

Several age clocks based on different biological parameters have been proposed ^38–40,42^ and a recent study developed an inflammatory clock (iAge) based on patterns of systemic age-related inflammation tagged to multimorbidity and frailty in the elderly ^36^. The strongest contributor to iAge was CXCL9, a chemokine involved in cardiac aging and impaired vascular function. Interestingly, CXCL9 is analogous to and from the same family of CXCL10, a mediator highly associated with severity of COVID-19 ^51^ (**Figures 1 and 2**). CXCL9 was found at high levels in hospitalized (**Figure 2d**), severely diseased (**Figure 2e, f**) patients and in the elderly from Belo Horizonte and Governador Valadares regardless of their infection status (**Figure 2d**). Therefore, at early stage of infection, severe COVID-19 could be distinguished from other clinical forms of the disease by a strong biomarker of aging.

Alongside with the inflammaging phenomena, immunosenscence is marked by several changes in innate and acquired immunity being lymphocytes and the T cell compartment the most affect ones. During aging, thymic atrophy, sustained replicative pressure due to homeostatic proliferation and clonal expansion after antigenic stimulation leads to the emergence of terminally differentiated T cells such as exhausted and senescent cells. Although these T cell populations share common features such as functional remodelling and cell cycle arrest, they have also distinctive characteristics ^52^. Chronically stimulated T cells during aging, persistent viral infections and cancer trigger an exhausted profile characterized by the expression of inhibitory receptors such as programmed cell death protein 1 (PD-1) ^53,54^, inducible costimulatory molecule (ICOS) ^53,54^ and T-cell immunoglobulin and immunoreceptor tyrosine-based inhibitory motif domain (TIGIT) ^55^. Exhausted T cells display a dysfunctional and non-proliferative state that is reversible. On the other hand, immune and non-immune senescent cells present a non-reversible loss of their proliferative capacity and acquire a senescence associated secretory phenotype (SASP) that contributes to inflammaging ^56^. Expression of CD57 ^56^ and killer cell lectin-like receptor G subfamily member 1 (KLRG1) ^56^ as well as the loss of the co-stimulatory molecule CD28^56^ are considered indicators of terminal differentiation and are associated with the senescence profile of T lymphocytes. Although KLRG1 also emerges on the cell surface after differentiation, the signalling generated by this molecule regulates a pathway related to exhaustion, making it a marker of both cellular senescence and exhaustion ^56^.

Notably, we also detected, in patients with severe COVID-19, an accrual of CD8^+^ T cells expressing markers of senescence (TIGIT) and exhaustion (ICOS) (**Figure 3a, b**) as well as increase in frequencies of senescent CD4^+^TIGIT^+^ T cells. Using CD28, KLRG1, CD57, TIGIT, PD-1 and ICOS as biomarkers, higher frequencies of CD8^+^ and CD4^+^ T cells with mixed senescence/ exhaustion phenotypes were also observed individuals with severe COVID-19 when compared to individuals with mild disease and, for specific phenotypes, to individuals with moderate disease (**Figure 3**). This association of T cell senescence to COVID-19 severity further confirmed our hypothesis and prompt us to examine other parameters of aging as risk factors for the worse disease outcomes.

To further understand the influence of B cell senescence in SARS-CoV-2 infection, we explored the immunoglobulin repertoire of COVID-19 patients and healthy individuals. Aging is associated with decreased lymphoid differentiation and with a consequent reduction in the output of naïve B cells. Clonal expansion of memory-like B cells in the periphery due to homeostatic proliferation and chronic stimulation by antigens result in maturation and shrinkage of the immunoglobulin repertoire ^7,8^. Herein, we analyzed the most expanded clones to have a better grasp of their representation in the overall repertoire. As shown in **Figure 4a**, the most expanded clones were not as dominating in individuals who tested negative for the disease when compared to the one who had a mild outcome. Also, we observed that individuals who were hospitalized had the most expanded clones representing a larger amount of the repertoire. In addition, hospitalized patients had lower repertoire diversity when compared to both negative and mild COVID-19 individuals (**Figure 4b**). These results correlated with previous reports demonstrating that the more severe the COVID-19 the lower the diversity of the antibody repertoire ^57,58^. In addition, the immunoglobulin repertoire of individuals who needed to be hospitalized had a more mature profile (**Figure 4c**) characterized by altered immunoglobulin gene usage and an increased frequency of mutated antibodies structurally diverging from their germline precursors. Mature repertoires are indicators of immunosenescence because mutations accumulated with age as the frequency of germline-encoded antibodies decrease ^8^.

To further explore the idea that individuals who developed severe COVID-19 might have an immunosenescent phenotype, we sought to investigate the biological age of individuals within our cohort measuring DNA methylation in CpG dinucleotides. Many studies have used DNA methylation as an epigenetic clock to measure biological age since DNA methylation accumulates with aging ^38^. The epigenetic age can be accelerated by certain diseases including HIV infection, Alzheimer’s, obesity, and by chronic exposure to infectious agents even when the individuals are not infected^42,16^.

To increase the accuracy in measuring biological age by an epigenetic approach, we analyzed the DNA methylation data using 8 different clocks: the classic and updated Horvarth clocks ^59^, Hannum’s, Levine’s, the BLUP, Wu, EN and TL clocks, the latter focused on CpGs associated with telomere formation, expression, and maintenance^27,43,46^. Our results show an acceleration of epigenetic age in hospitalized patients with severe COVID-19 in 7 of the 8 analyzed clocks if we exclude individuals from Governador Valadares. Including this population, we found a similar profile result (**Figure 4d**) with a variation in one of the clocks. However, there is an increase in statistical significance in the analysis when individuals from the endemic area were included (**Figure 4e**). Thus, these data confirmed, by another approach, the tight association between the immunosenescence phenotype and COVID-19 severity. Moreover, a network analysis of integrated data on the immunosenescence profile showed that individuals who developed severe COVID-19 had clusters with higher degree of integration mostly among senescent and exhausted of T cells (**Figure 4 f**). A graphical representation of all data illustrates that individuals with severe disease resemble the aged phenotype (**Figure 4 g**).

There were limitations in this study. Although we included only individuals with initial infection (1-4 days of symptoms), we cannot exclude the overlap between the inflammatory mediators coming from a previous state of inflammaging from the early innate response to SARS-CoV-2. This was less relevant for the analysis of senescent and exhausted T cells because we have shown that COVID-19 can induce senescence and exhaustion in CD8^+^ and CD4^+^ T cells after 7 days of symptoms ^27^. It is probably the case of the repertoire analysis of antibodies that are very unlikely to have expanded due to virus stimulation in such short time of infection. However, the choice of working with individuals within 1-4 days of symptoms and the need to match all the individuals by sex, age and presence of comorbidities reduced our study sample, which was another limitation.

Despite these restrictions, our data clearly show a close association between a profile of immunosenescence, examined by different approaches and consolidated by network analysis, and the progression of early phase infected individuals to severe COVID-19. We also reached important non-anticipated conclusions such as the uniqueness of the inflammatory pattern associated with early COVID-19 when compared to other respiratory infections and the differences in the inflammaging and immunesenescence phenotype in endemic regions for infectious disease when compared other metropolitan areas. These results are relevant not only to uncover the pathogenesis of severe COVID-19, but they have also predictive value for future epidemic viral infections. Most of all, the data shed light on the aging process and on the consequences of accelerated senescence that affect certain endemic regions of the planet.

## Author contribution

A.M.C. designed the study, coordinate the recruitment and analysis, and wrote the manuscript; L.H.A.V. and L.T. performed sample collection and processing, Luminex and flow cytometry analysis, and wrote the manuscript; G.C.C. performed the recruitment, blood collection and sample processing of the volunteers in Belo Horizonte, discussed the results and wrote the manuscript; J.Z. performed flow cytometry analysis of samples from São Paulo and discussed the results, M.M.C. performed DNA methylation analysis and discussed the results; C.H.R-P. was in the charge of sample processing and helped with the flow cytometry analysis, J.G. performed the B cell repertoire analysis and discussion under the supervision of L.F.F., F.C.M. performed the bioinformatic analysis and discussed the results; L.N. developed the database for the study, M.A.O., V.D.M. and M.F.O. helped in sample processing, Luminex analysis and discussion of the results, L.W.Z. helped with B cell repertoire and DNA methylation analysis and the discussion of the results; M.S.C. performed the clinical exams of volunteers at UPA-Centro Sul in Belo Horizonte under the supervision of U.T.; H.I.S. performed the RT-PCR tests of all volunteers under the supervision of S.M.R.T.; H.C.G. and R.C.B. were in charge of clinical exams of volunteers recruited at Hospital Universitário Risoleta Tolentino Neves in Belo Horizonte; A.P.V., N.A., G.P.C. and S.R. were in charge of clinical exams of volunteers recruited at Instituto de Infectologia Emílio Ribas, M.L.O.J. was in the charge of the clinical exams of volunteers recruited at Hospital Unimed in Governador Valadares; A.P.C.B. helped in the recruitment and sample processing of volunteers in Governador Valadares; F.F.C. helped in recruitment, sample collection and processing in Belo Horizonte; E.S.F. helped with the Luminex testing and data analysis supervised by O.A.M.F.; V.C. helped in the bioinformatic and statistical analysis as well as the discussion of the results, R.A. was in the charge of radiographic classification and follow-up of hospitalized patients in São Paulo; R.S. helped in the recruitment of healthy volunteers in São Paulo; G.S-N. coordinate all the recruitment and sample collection/processing of volunteers from Governador Valadares and helped in the statistical analysis; M.A. and C.L. helped in sample processing and the flow cytometry analysis of volunteers from São Paulo, T.U.M. coordinate the recruitment, questionnaire application and telemonitoring of volunteers of Belo Horizonte and helped discussing the data; D.M.F. coordinate sample processing and flow cytometry analysis in São Paulo, helped designing the experiments and discussing the data; A.T-C. supervised the flow cytometry analysis, Luminex analysis, serology tests in Belo Horizonte, helped designing the experiments and discussing the data.

## Acknowledgments

This study was supported by grants from Merck Dohme & Sharp (MISP#60383), Conselho Nacional de Desenvolvimento Científico e Tecnológico (CNPq, Brazil, 407363/2021-1), Coordenação de Aperfeiçoamento de Pessoal do Ensino Superior (# 0688/2020, CAPES, Brazil) and Pro-reitoria de Pesquisa da Universidade Federal de Minas Gerais (PRPq-UFMG). A.M.C.F., A.T-C., D.M.F., T.U.C., O.A.M.F., G.C.C., M.M.C., M.A.O., M.F.O., V.D.M. are recipients of research fellowships and scholarships from CNPq, L.H.A.V., L.T., L.N. are recipients of scholarships and fellowships from CAPES, Brazil, F.C.M. is recipient of a scholarship from Fundação de Amparo à Pesquisa do Estado de Minas Gerais (FAPEMIG, Brazil), M.A. and C.L. are recipients of a scholarships from Fundação de Amparo à Pesquisa do Estado de São Paulo (FAPESP, Brazil).

## Data availability statement

The MiAIRR data that support the findings of this study are available in Zenodo with the identifier(s) doi:10.5281/zenodo.12789370. FASTQ files can be accessed via NCBI under the BioProject accession PRJNA1138747 or SRA SRP528303, in the project Immunosenescence of antibody repertoire in individuals from endemic areas for infectious diseases or via the link: https://www.ncbi.nlm.nih.gov/bioproject/PRJNA1138747.

Data are also available upon request to the corresponding author.

## Materials and Methods

### Ethics Statement

The present study was approved by the national research ethics committee in Brazil (CONEP) and by the research ethics committees of the Universidade Federal de Minas Gerais (UFMG), and of the Instituto de Infectologia Emílio Ribas (IIER) (CAAE 40208320.3.2001.0061). All participants signed the informed consent form during the first evaluation, agreeing to participate in the study.

### Study Population and Data Collection

A total study sample of 806 volunteers were recruited in 3 cities, Belo Horizonte/MG (467 individuals), Governador Valadares/MG (176 individuals) and São Paulo/SP (163 individuals) from December 2020 to October 2021 (**Table 1 and 2**). The strains that circulated during the period were the original and P1 (Brazilian variant). Individuals with symptoms of a flu-like syndrome of unknown ethiology and healthy individuals were used as study controls. Volunteer recruitment in Belo Horizonte (MG) was carried out at the Health Care Unit in the Center-South region (UPA-CS), at Hospital Risoleta Tolentino Neves, at Casa do Ancião in Cidade Ozanam and at home. In São Paulo (SP), volunteer recruitment and sample collection took place at the Instituto de Infectologia Emílio Ribas (IIER) and in Governador Valadares (MG), at Hospital da Unimed de Governador Valadares and at home for some exceptional cases. A study flowchart with an outline of recruitment and sample collection at the health units is shown in **Supplementary Figure 4**. All individuals had their blood collected and were tested using nasopharyngeal swabs samples for reverse-transcriptase quantitative polymerase-chain-reaction (RT-qPCR) detection of SARS-CoV-2. They were telemonitored for 14 days to obtain information on their clinical status. Hospitalized individuals had their clinical evolution information collected directly from their medical records. All volunteers had their weight and height measured to determine their nutritional status by using the Body Mass Index (BMI) ratio. Weight was assessed using a Balmark® device with a capacity of 200 kg and an accuracy of 100 g, while height was measured with a portable stadiometer that accommodates up to 2 meters with an accuracy of 1.0 mm. The Body Mass Index (BMI) was calculated using the formula: BMI = weight (kg) / height² (m) and was subsequently classified according to their age range, based on the World Health Organization scale ^60^. A clinical and sociodemographic questionnaire was also administered during recruitment to gather additional information, such as current symptoms, days since symptom onset, vaccination status, and medical history.

### Study inclusion and exclusion criteria

Initially, the inclusion criteria for the study groups were all individuals over 20 years of age who had flu-like symptoms at the time of data collection. For negative individuals, the inclusion criterion at recruitment was all individuals who were never diagnosed with COVID-19 nor had flu-like symptoms with a confirmed negative RT-PCR test for SARS-CoV-2 infection and who tested negative for serological quantification of the following 4 viral proteins: spike 1, spike 2, nucleocapsid and receptor binding domain (RBD).

Exclusion criteria were applied to any patient who took any dose of the COVID-19 vaccine, patients living with HIV, those with inconclusive RT-PCR test, missing important information in their medical records or in the health questionnaire, those with greater than 4 days of symptom onset and those with a positive anti-SARS-CoV-2 IgG or IgM serology. We included only individuals who had 1 to 4 days of symptoms to better assess possible predictors of the disease. As shown previously by our group, individuals with mild and severe COVID-19 presented an increase in the frequency of exhausted/senescent T cells and in the levels of inflammatory mediators after 7 days of symptoms with a clear difference between the ones with only 4 days of symptom onset^27^.

### Clinical Groups and Criteria for Classification of COVID-19 Patients

We classified those infected with COVID-19 (positive RT-PCR for SARS-CoV-2) into different clinical forms of COVID-19 (mild, moderate and severe) according to WHO criteria (**Supplementary Table 1**). To perform certain analysis, we chose to group individuals with a moderate or severe clinical form of COVID-19 in a single group called Hospitalized since these individuals needed inpatient care. Individuals with flu-like symptoms who presented a negative RT-PCR and serological test for SARS-CoV-2 were grouped in the flu-like syndrome (FLS) group. A group of negative individuals was used in some analysis, and it consists of individuals with a negative SARS-CoV-2 test and no report of flu-like symptoms.

### RT-PCR for SARS-CoV-2 detection and viral load calculation

RNA extraction from nasopharyngeal swab samples was performed using the QIAamp Viral RNA Mini Kit (Qiagen, Germany) according to the manufacturer’s protocol. From 1 mL of Viral Transport Medium containing the collected sample, 47 µL of each individual sample was pooled in 1.5 mL microtubes, and 150 µL of the pool was used for extraction; the remainder was stored at −80 °C for possible repetitions. In cases where the clustering resulted in detectable results, it was necessary to individually process all samples present in the respective cluster. RT-qPCR was performed using the QuantStudio™ 3 and 5 real-time PCR systems (Applied Biosystems™, United States). For viral RNA detection, the recommendations described by the Charité protocol ^61^ were followed, which targets the gene (E) encoding the viral envelope protein, with a reported sensitivity of 3.9 copies of the SARS-CoV-2 genome per reaction ^61^. Thermal cycling was performed at 55°C for 10 min for reverse transcription, followed by 95°C for 3 min and then 45 cycles of 95°C for 15 s, 58°C for 30 s. For endogenous control of the reaction, the gene encoding human RNaseP was used as a target.

### Blood collection and isolation of PBMC

Peripheral blood mononuclear cells (PBMC) were obtained by blood collection from COVID-19 and control patients using heparinized vacuette. The purification was attained by Ficoll gradient (Histopaque – 1077; Sigma) centrifugation at room temperature, in a 1:2 ratio, for 40 minutes, at 1400 resolutions per minute (RPM) without breaking. After that, PBMCs were collected using a Pasteur pipette, washed with RPMI medium at 500x for 7 minutes at 4°C, and then red cells were lysed using a lysis buffer. A second wash using RPMI was conducted. Cells were counted and stored in fetal bovine serum supplemented with 10% dimethyl sulfoxide at −80°C until use. Samples were processed up to 48 hours after collection.

### Plasma and Serum Isolation

Blood was collected into heparin tubes for plasma obtention, and in serum separator tubes containing clot activator. Plasma and serum samples were isolated after whole blood centrifugation at 3000 RPMs for 10 minutes at 20°C, then carefully split into 2mL aliquots and stored at −80°C until the time of testing, Samples were processed up to 48 hours after collection.

### Measurement of cytokines, chemokines, and growth factors by Luminex-Multiplex

The measurement of biomarkers was performed in heparinized plasma for detection and quantification of analytes for all 309 individuals from Belo Horizonte, São Paulo and Governador Valadares (endemic area) all in Brazil. The Bio-Rad Laboratories kit (Bio-Plex® Pro Human Cytokine Standard) was used, allowing for the analysis of several analytes to be analyzed simultaneously, using the magnetic immunoassay technique carried out with the Luminex equipment (Bio-Plex® 200, Bio-Rad), following storage and processing protocols standardized by the Grupo Integrado de Pesquisa com Biomarcadores (GIPB) (IRR-FIOCRUZ/MG). Samples were transported and stored at a temperature of −80°C. Analyses were performed using Bioplex™ xPONENT version 3.1 software (Bio-Rad), and presented the following panel of analytes: IL-1β, IL-1Ra, IL-2, IL-4, IL-5, IL-6, IL-7, CXCL8, IL-9, IL-10, IL-12p70, IL-13, IL-15, IL-17A, CCL11, Basic-FGF, G-CSF, GM-CSF, IFN-γ, CXCL10, CCL2, CCL3, CCL4, PDGF-BB, CCL5, TNF and VEGF.

In all the inflammatory analysis carried out in the present study, we identified 10 of the 27 mediators evaluated as those that are most present in the analyses, configuring a possible signature of these mediators in our population (CCL2, CXCL8, CXCL10, IL-6, IL12(p70), IFN-γ, IL-1Ra, IL-10, TNF, IL-6) and in order not to dilute the data, we chose to show the differences only within this signature.

### Measurement of chemokines by Cytometric Bead Array (CBA)

The CBA assay was performed using the BD Cytometric Bead Array (CBA) – Human Chemokine Kit to complete the Luminex Bio-plex assay biomarker panel. The chemokines present in the kit are CCL2, CCL5, CXCL8, CXCL9 and CXCL10. Samples from Belo Horizonte-MG, Governador Valadares-MG and São Paulo-SP, all previously prepared before the test, were included. Kit reagents were previously filtered, and kit protocol was standardized and used. Samples were purchased from BD FACSVerse.

### Measurement of SARS-CoV-2 viral proteins by Luminex-Multiplex

The measurement of biomarkers was performed in serum for detection and quantification of proteins for 144 negatives individuals from Belo Horizonte, São Paulo and Governador Valadares (endemic area) all in Brazil. The Bio-Plex Multiplex SARS-CoV-2 Serology Assay Kit was used, allowing the analysis of the following viral proteins: Spike 1, Spike 2, Nucleocapsid and receptor binding domain (RBD). All samples were analyzed simultaneously using the magnetic immunoassay technique performed with the Luminex equipment (Bio-Plex® 200; Bio-Rad) and according to the storage and processing protocols standardized by the Grupo Integrado de Pesquisa com Biomarcadores (GIPB) (IRR - FIOCRUZ/MG). Samples were transported and stored at −80°C. Analysis were performed using Bioplex™ xPONENT software version 3.1 (Bio-Rad).

### Measurement of Vitamin D

The biochemical test to measure vitamin D levels was outsourced to a clinical analysis laboratory in Belo Horizonte, which has all the laboratory quality certifications in order. The serum levels of this vitamin were measured by chemiluminescence using the Atellica Siemens device. All tests were performed respecting the transport and storage conditions established by the outsourced company.

### Immunophenotype by polychromatic flow cytometry

PBMCs (1×10^6^) were first stained with LIVE/DEAD fixable aqua dead cell stain (ThermoFisher Scientifc, cat #L34957) and with fluorophore-conjugated monoclonal antibodies to human surface markers. All information on the antibodies, their concentrations, manufacturer and catalogue numbers can be found in **Supplemmentary Table 2**. After the surface staining, cells were fixed and permeabilized using the Foxp3/Transcription Factor Staining Buffer Set from eBioscience (cat #00-5523-00) and stained for -Foxp3 from ImmunoTools (3G3, cat# 21276106) (Supplemmentary Table 2). Single-color labelled cells for fluorescence compensation were prepared with antibody capture compensation beads (BD Biosciences). Cell samples were acquired in a BD LSRFortessa cell analyzer (BD Biosciences) coupled to computers with DIVA and FlowJo-10 *software* (Tree Star). One hundred events for each sample were acquired in the lymphocyte gate. The gate strategy used to analyze T lymphocyte populations is shown in **Supplementary Figure 3**.

### Extraction of DNA and RNA from PBMC and Bisulphite Treatment

Frozen samples of PBMCs were briefly thawed in a 37°C water bath and kept on ice. Pellets were then resuspended in 1ml Trizol and incubated at room temperature. Chloroform was then added to each sample. After homogenization and further incubation, samples were centrifuged to form a three-phase solution. The aqueous phase was used for RNA extraction and the interphase was used for DNA extraction. Thus, both samples underwent purification and quantification steps and, after elution, were stored in a −80°C freezer in low-binding plastic microtubes. Genomic DNA extraction was performed using the QIAamp 96 DNA Blood kit (QIAGEN, Hilden, Germany). DNA (1 μg) was bisulphite-converted using the EZ-96 DNA Methylation Kit (Zymo Research, Irvine, USA) with the following modifications: incubation in CT buffer for 21 cycles of 15 min at 55 °C and 30 s at 95 °C, elution of bisulphite-treated DNA in 100 μl of water. After extraction and conversion, samples were loaded on the Illumina Infinium MethylationEPIC BeadChip.

### Analyses of Epigenetic Aging: Obtaining Raw Methylation Data

DNA from the PBMC samples was extracted according to the Trizol and chloroform protocol. Subsequently, DNA was hybridized using the Illumina Infinium MethylationEPIC Beadchip (EPIC array) that covers nearly 850,000 CpG sites in the human genome. The watermelon 2.4.0 R package (Pidsley et al., 2013) was used to perform raw data processing, batch effect correction, and obtain matrices with methylation β value. The methylation β value for each CpG site in each sample was calculated to represent the level of methylation.

### DNA methylation age calculation

We used the mDNA age of epigenetic clocks created based on blood samples, including the Horvath clock with 353 CpGs based on various tissue types, the skinHorvath clock that integrated the whole blood dataset with skin, endothelial cells, buccal mucosa and saliva, the Levine clock based on 513 CpGs derived from whole blood, the TL telomere length clock based on 140 CpGs derived from blood, the BLUP clock based on 319607 CpGs derived from blood, the EN clock based on 514 CpGs derived from all blood and the Hannum watch with 71 CpGs identified in blood DNA samples. The deviation between epigenetic age and chronological age, also known as epigenetic age acceleration, was calculated for each sample based on mDNA age regression residuals considering cell counts with the R methylclock 1.5.0 package ^62^.

### B cells repertoire analysis using immunoglobulin variable heavy chain (IGVH) amplification

A subset of 45 individuals was analyzed to examine their B cell repertoire. For this, RNA was extracted from frozen PBMC samples of these individuals using Trizol (ThermoFisher). The cDNA was synthesized with SuperScript IV First-Strand Synthesis System (ThermoFisher) from approximately 500 ng of RNA, following the manufacturer’s instructions. IGVH amplification was performed using 0.2 mM of forward primers (1 to 8; **Supplementary Table 4**), 0.1 mM of each IgG and IgA reverse primers (9 and 10; Supplementary Table 4), 0.2 units (U) of Platinum™ Taq DNA Polymerase High Fidelity (Invitrogen), 1X High Fidelity PCR Buffer solution, 0.2 mM of dNTPs, 1 mM of MgSO_4_, and 2 µL of cDNA in a final volume of 50 µL. The PCR reaction was conducted using the following program: 2 minutes (min) at 95 °C; 4 cycles of 94°C for 30 seconds (sec), 50°C for 30 sec, and 68°C for 1 min; 4 cycles of 94°C for 30 sec, 55°C for 30 sec, and 68°C for 1 min; 22 cycles of 94°C for 30 sec, 63°C for 30 sec; 68°C for 7 min and final hold at 4°C. Samples GV47, GV50, GV51, GV54, GV92, GV106, GV144, and GV146 were amplified using the NEBNext® High-Fidelity 2X PCR Master Mix (New England Biosciences, UK). Forward primers at a final concentration of 0.2 mM, IgG/IgA reverse primers at 0.1 mM, and 2 µL of cDNA were added to 1x PCR Master Mix in a final volume of 50 µL. The cycling conditions were the same as described above, except for an extension temperature of 72° C. Four replicates were amplified for each sample. The mixed 200 µL of PCR product of each sample was precipitated using Puregene® Concentrating DNA protocol (Qiagen, Germany). IgG/IgA amplification was excised from an electrophoresis run of 1% agarose gel, followed by the PCR purification protocol provided by NucleoSpin Gel and PCR Clean[up kit (Macherey-Nagel). The concentration of the DNA was calculated using Nanodrop and Qubit DNA High Sensitivity kit (ThermoFisher Scientific).

### Library construction and sequencing

Sequencing libraries were constructed using the Nextera XT DNA Library Prep Kit (Illumina) according to the manufacturer’s instructions. A PCR reaction was prepared to contain: 5 µl of Nextera CD Index 1 (i7) Primer (H7XX); 5 µl of Nextera CD Index 2 (i5) Primer (H5XX); 1X of the High-Fidelity PCR Buffer solution; 0,2 mM of dNTPs; 1 mM of MgSO_4_; 1 U of Platinum® Taq DNA High Fidelity Polymerase; 5 µl (if DNA concentration >= 10 ng/µl) or 10 µl (if DNA concentration ranged from 5-10 ng/µl) of the gel purified amplicon in a final volume of 50 µl. The cycling condition was 3 min at 95 °C; 12 cycles of 94°C for 30 sec, 55°C for 30 sec, and 68°C for 1 min; 68°C for 5 min; and hold at 4°C. The PCR product was purified with 0.5 volumes of Agencourt AMPure XP beads and the library was quantified using the Qubit DNA High Sensitivity kit (ThermoFisher Scientific). Amplicon size was inferred using the High Sensitivity DNA Kit (Agilent) run on the Bioanalyzer 2100 (Agilent). Sequencing was performed on the Illumina MiSeq System (Illumina), using the Miseq Reagent Kit v3 (600-Cycle) (Illumina) with 301 sequencing cycles for each forward and reverse read. A total of 31 different libraries from the volunteers were joined equimolarly in a pool of 4nM and diluted to 18 picomolar for further sequencing. A summary of found sequences are in **Supplementary Table 4.**

### Bioinformatic analysis: Pre-processing

Samples were demultiplexed by the Illumina sequencer tool or using an in-house script resulting in FASTQ files. All pre-processing was done using the framework Immcantation and it consists of 4 steps: 1) Using pRESTO ^63^, the AssemblePairs module was used to join the sequence, based on the 3’-5’ read and the 5’-3’ read, with a minimum overlap of 50 nucleotides; 2) Quality filter was applied by using the module FilterSeq ^63^ Only sequences with a Phred score >=30 were selected; 3) The module MaskPrimers ^63^ was employed to find the primers of the IGHV gene and constant region. The option score was employed, preserving sequences with both forward and reverse primers intact; any sequences not meeting these conditions were filtered out; 4) Annotation was performed with the AssignGenes module from Change-O ^63^ together with IgBlast ^64^ to verify similarities between the subject and query. IMGT human germline was used as a reference for V, J, and D genes, as well as their numbering system ^65^; the MakeDB module was run to organize the alignment into a tabular .tsv file in the MiAIRR format ^66^. This format contains information on V, D, and J genes and alleles, framework (fw), complementary regions sequences (cdr), and functional status of the sequence (productive or unproductive).

### Bioinformatic analysis: Clonotyping and Diversity Analysis

The sequences were grouped into clones using the tool YClon ^67^. Shannon diversity was calculated using the *diversity* function of the R package ‘vegan’ (https://CRAN.R-project.org/package=vegan). Diversity was compared between groups of individuals with flu-like symptoms or control (*n*=6), with the ones with mild COVID (n=19), and severe COVID-19 (n=20) from Belo Horizonte, São Paulo and Governador Valadares (only mild cases). Diversity was computed for the 100 most expanded clones from each repertoire. The complete pipeline employed to prepare and analyze the IGH repertoire is available on https://github.com/jao321/covidao.git.

### Repertoire Maturity Analysis

We define repertoire maturity as the extent to which an individual’s immune repertoire has diverged from germline-like sequences through processes such as somatic hypermutation and clonal selection. A more mature repertoire is characterized by a lower proportion of germline-like clones, reflecting immune system adaptation and experience with antigens over time. To gain a more profound insight into the maturity of the repertoire, an investigation was conducted to identify the sequence closest to the germline for each clone, encompassing both the V and J genes. To achieve this, we considered the difference in length between the V gene (5 to 6 times longer than the J gene) to ensure a comprehensive analysis, using the following calculation:

VJpid =(Vpid ∗0.8)+(Jpid ∗0.2)

VJpid: Percentage of the expected identity of the V and J genes when compared to the germline

VJpid: Percentage of identity of V genes compared to germline

VJpid: Percentage of the identity of the J gene when compared to the germline

Only the sequence with the highest VJpid value for each clone remained, as these sequences have undergone fewer mutations.

### Statistical Analysis

The Shapiro-Wilk and Kolmogorov-Smirnov tests were applied to determine whether the data had a normal (parametric) or non-normal (non-parametric) distribution. Outliers were excluded using the ROUT test. For each group, Mann-Whitney test or Kruskal-Wallis with Dunn’s post-test was performed depending on the data distribution normal or non-normal. The violin plot was chosen to demonstrate the statistical significance of our analyses. Its format is directly proportional to the dispersion of the data, the dotted lines correspond to the minimum quartiles in the lower part and maximum in the upper part, and the solid line represents the median. Data are represented by black dots (each dot represents an individual) and were normalized in log10. In addition, all groups received a specific color that represents them in all graphs and figures.

### Radar-Chart

The radar chart is a graph that represents the frequency (in percentage) of high producers of each plasma mediator represented and it has been previously described ^17^. On the charts, each axis represents the percentage (%) of volunteers showing high levels of a specific mediator. The values of each axis can be connected to form a central polygonal area that represents 50% of the baseline level and the external polygonal area is equivalent to 100%. Increase or decrease of the central polygonal area reflects either a higher or a lower contribution of the mediators for each group. We consider all mediators that are above the internal polygonal area to be high producers.

### Network analysis

A weighted network analysis was used to investigate the correlations between biologically relevant markers associated with cytokines and cellular proteins (listed in **Supplementary Table 5** and selected based on their potential relevance for the study). To quantify the relationships between these markers, we utilized Spearman Rank correlation, a non-parametric method that ranks the data points for each variable and calculates the correlation coefficient based on the ranks. This approach is suitable for data that may not follow a normal distribution or have outliers. The correlation coefficient ranges from −1 to 1, where 1 indicates a perfect positive correlation, −1 indicates a perfect negative correlation, and 0 indicates no correlation. To focus on significant correlations, we established a threshold of p-values less than 0.05. This filtering step ensured that only statistically significant relationships were included in the network analysis. The resulting network was visualized using a spring layout, a graph drawing algorithm that positions nodes based on the forces of attraction and repulsion between them. This layout is particularly effective for identifying clusters or modules within the network, providing insights into the underlying structure of the relationships. The strength of correlations was visually represented by the darker of the lines connecting nodes. Darker lines indicated stronger relationships between variables, while lighter lines suggested weaker associations. Additionally, the size of each node corresponded to the number of correlations it participated in, providing a visual cue for the variable’s centrality within the network. Larger nodes represented variables with more connections, highlighting their importance in the overall system. By visualizing the network in this manner, we were able to gain insights into the interactions between cytokines/chemokines and cellular proteins, identifying key players and potential regulatory mechanisms within the biological system under study.

