## Supplementary Tables for "Immunosenescence profile is associated with increased susceptibility to severe COVID-19"

**Suplemmentary Table 1: Who criteria for the clinical groups**

| *Patient State* | *Descriptor* | *Score* |
|---|---|---|
| *Uninfected* | Uninfected; no viral RNA detected. | 0 |
| *COVID-19 Mild* | Assymptomatic; viral RNA detected. | 1 |
| | Symptomatic; Independent. | 2 |
| | Symptomatic; Assistance needed. | 3 |
| *COVID-19 Moderate* | Hospitalised; No oxygen therapy. | 4 |
| | Hospitalised; Oxygen by mask or nasal prongs. | 5 |
| *COVID-19 Severe* | Hospitalised; Oxygen by NIV or high flow. | 6 |
| | Intubation and mechanical ventilation, $pO_2/FiO_2 \geq 150$ or $SpO_2/FIO_2 \geq 200$). | 7 |
| | Mechanical ventilation, $pO_2/FiO_2 < 200$) or vasopressor. | 8 |
| | Mechanical ventilation, $pO_2/FiO_2 < 150$ and vasopressor, dyalysis or ECMO. | 9 |
| | Dead. | 10 |

**Suplemmentary Table 2: Panel of mAbs for flow cytometry**

| | Antibody | Channel | Dilution Used | Catalog Number | Clone |
|---|---|---|---|---|---|
| T Cell Panel | CD4 | Pe Cy 7 | 1:400 | 557852 | SK3 |
| | CD8 | PE-Dazzle 594 | 1:1200 | 562282 | RPA-T8 |
| | CD25 | PE | 1:20 | 555432 | M-A251 |
| | CD278 (ICOS) | Percp Cy 5.5 | 1:20 | 562833 | DX29 |
| | CD57 | BV 421 | 1:800 | 563896 | NK-1 |
| | CD279 (PD-1) | BV 605 | 1:40 | 563245 | EH12.1 |
| | TIGIT | BV 650 | 1:160 | 741182 | FAB7898G |
| | CD28 | APC H7 | 1:20 | 561368 | CD28.2 |
| | CD3 | AF 700 | 1:320 | 317340 | OKT3 |
| | CCR7 | FITC | 1:40 | 353216 | G943H7 |
| | KLRG-1 | BV 711 | 1:40 | 138427 | 2F1/KLRG1 |
| | FOXP3 | APC Cy7 | 1:20 | 21276106 | 3G3 |
| | CD45RO | BV 785 | 1:80 | 304234 | UCHL1 |
| NK Cell Panel | CD3 | AF 700 | 1:320 | 317340 | OKT3 |
| | CD16 | PE Cy 5.5 | 1:100 | 555408 | 3G8 |
| | CD25 | PE | 1:20 | 555432 | M-A251 |
| | CD56 | PE Cy 7 | 1:200 | 340723 | NCAM 16.2 |
| | CD57 | BV 421 | 1:640 | 563896 | NK-1 |
| | CD69 | APC Cy7 | 1:20 | 557756 | FN50 |
| | CD 279 (PD-1) | BV 605 | 1:40 | 563245 | EH12.1 |
| | NKG2D | APC | 1:20 | 558071 | 1D11 |
| B Cell Panel | CD24 | FITC | 1:160 | 555427 | ML5 |
| | CD 279 (PD-1) | BV 605 | 1:40 | 563245 | EH12.1 |
| | CD38 | PE | 1:20 | 555460 | HIT2 |
| | CD5 | Percp Cy 5.5 | 1:80 | 341089 | L17F12 |
| | CD19 | PE Cy 7 | 1:640 | 557835 | SJ25C1 |
| | IgD | AF 700 | 1:40 | 561302 | IA6-2 |
| | CD138 | APC | 1:20 | 347193 | MI15 |
| | CD27 | APC Cy7 | 1:20 | 560222 | M-T271 |
| | CD20 | BV 650 | 1:800 | 563780 | 2H7 |

**Suplemmentary Table 3: Panel of mAbs for flow cytometry**

| City | Clinical Outcome | Sample | Raw read | Pre-processed reads* | Annotated reads | Clones |
|---|---|---|---|---|---|---|
| **Belo Horizonte** | **Control** | **A04** | 406,894 | 246,973 | 244,359 | 16,300 |
| **Belo Horizonte** | **Control** | **A20** | 618,809 | 420,539 | 418,354 | 12,662 |
| **Belo Horizonte** | **Control** | **A24** | 1,362,447 | 411,169 | 403,966 | 9,567 |
| **Belo Horizonte** | **Control** | **A65** | 555,155 | 348,297 | 344,143 | 15,331 |
| **Belo Horizonte** | **Control** | **A66** | 761,554 | 478,945 | 472,011 | 17,086 |
| **Belo Horizonte** | **Control** | **A67** | 546,518 | 354,867 | 350,617 | 31,830 |
| **Governador Valadares** | **Mild** | **GV106** | 537,812 | 255,051 | 199,460 | 6,956 |
| **Governador Valadares** | **Mild** | **GV144** | 349,768 | 215,476 | 193,437 | 9,710 |
| **Governador Valadares** | **Mild** | **GV146** | 1,450,041 | 398,780 | 395,181 | 9,087 |
| **Governador Valadares** | **Mild** | **GV43** | 906,435 | 279,639 | 271,681 | 13,703 |
| **Governador Valadares** | **Mild** | **GV47** | 544,092 | 202,199 | 159,930 | 6,907 |
| **Governador Valadares** | **Mild** | **GV50** | 1,332,292 | 299,198 | 236,633 | 8,494 |
| **Governador Valadares** | **Mild** | **GV51** | 1,477,827 | 306,737 | 295,683 | 8,186 |
| **Governador Valadares** | **Mild** | **GV54** | 628,211 | 264,316 | 245,602 | 8,940 |
| **Governador Valadares** | **Mild** | **GV68** | 558,832 | 344,804 | 338,562 | 13,168 |
| **Governador Valadares** | **Mild** | **GV92** | 487,172 | 203,943 | 130,060 | 3,053 |
| **Belo Horizonte** | **Mild** | **ID094** | 620,176 | 410,089 | 408,93 | 7,021 |
| **Belo Horizonte** | **Mild** | **ID117** | 540,888 | 341,650 | 340,256 | 13,239 |
| **Belo Horizonte** | **Mild** | **ID124** | 632,642 | 427,965 | 422,618 | 36,806 |
| **Belo Horizonte** | **Mild** | **ID131** | 536,784 | 311,349 | 304,709 | 9,865 |
| **Belo Horizonte** | **Mild** | **ID132** | 436,757 | 264,768 | 262,704 | 32,069 |
| **Belo Horizonte** | **Hospitalized** | **ID141** | 412,936 | 235,970 | 233,636 | 10,807 |
| **Belo Horizonte** | **Hospitalized** | **ID143** | 341,073 | 232,296 | 231,034 | 11,192 |
| **Belo Horizonte** | **Hospitalized** | **ID144** | 510,649 | 347,086 | 337,768 | 9,042 |
| **Belo Horizonte** | **Mild** | **ID155** | 557,750 | 323,370 | 311,478 | 14,827 |
| **Belo Horizonte** | **Hospitalized** | **ID187** | 563,901 | 295,370 | 284,909 | 26,310 |
| **Belo Horizonte** | **Hospitalized** | **ID195** | 406167 | 233,411 | 229,688 | 6,807 |
| **Belo Horizonte** | **Hospitalized** | **ID226** | 1,525,890 | 333,602 | 330,351 | 15,422 |
| **Belo Horizonte** | **Mild** | **ID240** | 407,527 | 236,377 | 229,633 | 8,234 |
| **Belo Horizonte** | **Mild** | **ID244** | 430,185 | 260,499 | 256,455 | 18,387 |
| **Belo Horizonte** | **Hospitalized** | **ID248** | 405,371 | 277,585 | 272,233 | 18,185 |
| **Belo Horizonte** | **Hospitalized** | **ID268** | 285,341 | 180,243 | 179,320 | 30,896 |
| **Belo Horizonte** | **Hospitalized** | **ID310** | 638,973 | 418,567 | 417,669 | 14,627 |
| **Belo Horizonte** | **Hospitalized** | **ID375** | 638,037 | 379,779 | 374,808 | 69,737 |
| **São Paulo** | **Hospitalized** | **SP10** | 386,469 | 205,262 | 194,655 | 23,650 |
| **São Paulo** | **Hospitalized** | **SP114** | 429,407 | 243,112 | 238,327 | 9,007 |
| **São Paulo** | **Hospitalized** | **SP138** | 790,021 | 550,456 | 548,060 | 23,304 |
| **São Paulo** | **Mild** | **SP14** | 521,830 | 324,709 | 321,397 | 41,497 |
| **São Paulo** | **Hospitalized** | **SP20** | 639,344 | 366,906 | 363,329 | 12,671 |

*Reads after steps 1, 2, and 3 of pre-processing.

**Suplemmentary Table 4: Summary of VIg sequences (repertoire)**

| | NAME | SEQUENCE (5' TO 3') |
|---|---|---|
| **1** | VH1-fwd | **TCGTCGGCAGCGTCAGATGTGTATAAGAGACAG**CAGGTCCAGCTKGTRCAGTCTGG |
| **2** | VH157-fwd | **TCGTCGGCAGCGTCAGATGTGTATAAGAGACAG**CAGGTGCAGCTGGTGSARTCTGG |
| **3** | VH2-fwd | **TCGTCGGCAGCGTCAGATGTGTATAAGAGACAG**CAGRTCACCTTGAAGGAGTCTG |
| **4** | VH3-fwd | **TCGTCGGCAGCGTCAGATGTGTATAAGAGACAG**GAGGTGCAGCTGKTGGAGWCY |
| **5** | VH4-fwd | **TCGTCGGCAGCGTCAGATGTGTATAAGAGACAG**CAGGTGCAGCTGCAGGAGTCSG |
| **6** | VH4-DP63-fwd | **TCGTCGGCAGCGTCAGATGTGTATAAGAGACAG**CAGGTGCAGCTACAGCAGTGGG |
| **7** | VH6-fwd | **TCGTCGGCAGCGTCAGATGTGTATAAGAGACAG**CAGGTACAGCTGCAGCAGTCA |
| **8** | VH3N-fwd | **TCGTCGGCAGCGTCAGATGTGTATAAGAGACAG**TCAACACAACGGTTCCCAGTTA |
| **9** | IgG-rev | **GTCTCGTGGGCTCGGAGATGTGTATAAGAGACAG**AGGGYGCCAGGGGGAAGAC |
| **10** | IgA-rev | **GTCTCGTGGGCTCGGAGATGTGTATAAGAGACAG**CGGGAAGACCTTGGGGCTGG |

The bold sequences constitute elements of the Illumina adaptor sequences. The design of the annealing region in primers was based on MacDaniel et al. (2016).

**Suplemmentary Table 5: Summary of Biomarkers of Network Analysis**

| ID | BIOMARKER |
|---|---|
| **1L** | CCL11 (Luminex) |
| **2L** | CXCL8 (Luminex) |
| **3L** | CCL2 (Luminex) |
| **4L** | CCL3 (Luminex) |
| **5L** | CCL4 (Luminex) |
| **6L** | CCL5 (Luminex) |
| **7L** | CXCL10 (Luminex) |
| **8PI** | IL-1$\beta$ (Luminex) |
| **9PI** | IL-6 (Luminex) |
| **10PI** | TNF (Luminex) |
| **11PI** | IL-12(p70) (Luminex) |
| **12PI** | IFN-$\gamma$ (Luminex) |
| **13PI** | IL-15 (Luminex) |
| **14PI** | 17-A (Luminex) |
| **15R** | IL-1Ra (Luminex) |
| **16R** | IL-4 (Luminex) |
| **17R** | IL-5 (Luminex) |
| **18R** | IL-9 (Luminex) |
| **19R** | IL-10 (Luminex) |
| **20R** | IL-13 (Luminex) |
| **21GF** | FGF-basic (Luminex) |
| **22GF** | VEGF (Luminex) |
| **23GF** | PDGF-BB (Luminex) |
| **24GF** | G-CSF (Luminex) |
| **25GF** | GM-CSF (Luminex) |
| **26GF** | IL-7 (Luminex) |
| **27GF** | IL-2 (Luminex) |
| **1C** | CCL2 (CBA) |
| **2C** | CCL5 (CBA) |
| **3C** | CXCL8 (CBA) |
| **4C** | CXCL9 (CBA) |
| **5C** | CXCL10 (CBA) |
| **AGE** | Age |
| **OUTCOME** | Outcome |
| **1T** | Total Cells |
| **2T** | Total Live Cells |
| **3T** | Lymphocytes |
| **4T** | Lymphocytes Live |
| **5T** | Lymphocytes CD3+ |
| **6T** | Lymphocytes CD3+CD4+ |
| **7T** | Lymphocytes CD3+CD4+CD25(High) |
| **8T** | Lymphocytes CD3+CD4+CD25(Low) |
| **9T** | Lymphocytes CD3+CD4+CD25+FOXP3+ |
| **10T** | Lymphocytes CD3+CD4+CD25+FOXP3+PD1+ |
| **11T** | Lymphocytes CD3+CD4+CD28-CD4+ |
| **12T** | Lymphocytes CD3+CD4+CD28-CD4+/CD4+CD28-CD57+ |
| **13T** | Lymphocytes CD3+CD4+CD28-CD4+/CD4+CD28-KLRG1+ |
| **14T** | Lymphocytes CD3+CD4+CD28-CD4+/CD4+CD28-PD-1+ |
| **15T** | Lymphocytes CD3+CD4+CD28-CD4+/CD4+CD28-TIGIT+ |
| **16T** | Lymphocytes CD3+CD4+CD28-CD4+/Q1:CD57-KLRG1+ |
| **17T** | Lymphocytes CD3+CD4+CD28-CD4+/Q2:CD57+KLRG1+ |
| **18T** | Lymphocytes CD3+CD4+CD28-CD4+/Q3:CD57+KLRG1- |
| **19T** | Lymphocytes CD3+CD4+CD28-CD4+/Q4:CD57-KLRG1- |
| **20T** | Lymphocytes CD3+CD4+/CD57+CD4+ |
| **21T** | Lymphocytes CD3+CD4+/CM CD4+ |
| **22T** | Lymphocytes CD3+CD4+/EFF CD4+ |
| **23T** | Lymphocytes CD3+CD4+/EFF CD4+/ CD4+ EFF CD28- |
| **24T** | Lymphocytes CD3+CD4+/EFF CD4+/ CD4+ EFF CD28-/ Q1: CD57-KLRG1+ |
| **25T** | Lymphocytes CD3+CD4+/EFF CD4+/ CD4+ EFF CD28-/ Q2: CD57+KLRG1+ |
| **26T** | Lymphocytes CD3+CD4+/EFF CD4+/ CD4+ EFF CD28-/ Q4: CD57+KLRG1- |
| **27T** | Lymphocytes CD3+CD4+/EFF CD4+/ CD4+ EFF CD28-/ Q4: CD57-KLRG1- |
| **28T** | Lymphocytes CD3+CD4+/EM CD4+ |

| | |
|---|---|
| **29T** | Lymphocytes CD3+CD4+/EM CD4+/ EM CD28- CD45RO+ |
| **30T** | Lymphocytes CD3+CD4+/EM CD4+/ EM CD28- CD45RO+/ CD4+ EM CD28-CD57+ |
| **31T** | Lymphocytes CD3+CD4+/EM CD4+/ EM CD28- CD45RO+/ CD4+ EM CD28-KLRG1+ |
| **32T** | Lymphocytes CD3+CD4+/EM CD4+/ EM CD28- CD45RO+/ CD4+ EM CD28- PD-1+ |
| **33T** | Lymphocytes CD3+CD4+/EM CD4+/ EM CD28- CD45RO+/ Q1: CD57-KLRG1+ |
| **34T** | Lymphocytes CD3+CD4+/EM CD4+/ EM CD28- CD45RO+/ Q2: CD57+KLRG1+ |
| **35T** | Lymphocytes CD3+CD4+/EM CD4+/ EM CD28- CD45RO+/ Q3: CD57+KLRG1- |
| **36T** | Lymphocytes CD3+CD4+/EM CD4+/ EM CD28- CD45RO+/ Q4: CD57-KLRG1- |
| **37T** | Lymphocytes CD3+CD4+/EM CD4+/ EM CD28- CD45RO+/ Q5: CD57-ICOS+ |
| **38T** | Lymphocytes CD3+CD4+/EM CD4+/ EM CD28- CD45RO+/ Q6: CD57+ICOS+ |
| **39T** | Lymphocytes CD3+CD4+/EM CD4+/ EM CD28- CD45RO+/ Q7: CD57+ICOS- |
| **40T** | Lymphocytes CD3+CD4+/EM CD4+/ EM CD28- CD45RO+/ Q8: CD57-ICOS- |
| **41T** | Lymphocytes CD3+CD4+/ KLRG1+CD4+ |
| **42T** | Lymphocytes CD3+CD4+/ NAÏVE CD4+ |
| **43T** | Lymphocytes CD3+CD4+/ PD-1+CD4+ |
| **44T** | Lymphocytes CD3+CD4+/ Q1: CD57- PD-1+ |
| **45T** | Lymphocytes CD3+CD4+/ Q2: CD57+ PD-1+ |
| **46T** | Lymphocytes CD3+CD4+/ Q3: CD57+ PD-1- |
| **47T** | Lymphocytes CD3+CD4+/ Q4: CD57- PD-1- |
| **48T** | Lymphocytes CD3+CD4+/ Q5: CD57- TIGIT+ |
| **49T** | Lymphocytes CD3+CD4+/ Q6: CD57+ TIGIT+ |
| **50T** | Lymphocytes CD3+CD4+/ Q7: CD57+ TIGIT- |
| **51T** | Lymphocytes CD3+CD4+/ Q8: CD57- TIGIT- |
| **52T** | Lymphocytes CD3+CD4+/ Q9: KLRG1- PD-1+ |
| **53T** | Lymphocytes CD3+CD4+/ Q10: KLRG1+ PD-1+ |
| **54T** | Lymphocytes CD3+CD4+/ Q11: KLRG1+ PD-1- |
| **55T** | Lymphocytes CD3+CD4+/ Q12: KLRG1- PD-1- |
| **56T** | Lymphocytes CD3+CD4+/ TIGIT+ CD4+ |
| **57T** | Lymphocytes CD3+CD8+ |
| **58T** | Lymphocytes CD3+CD8+/ CD8+CD28+ |
| **59T** | Lymphocytes CD3+CD8+/ CD8+CD28+/ CD8+CD28+CD57+ |
| **60T** | Lymphocytes CD3+CD8+/ CD8+CD28+/ CD8+CD28+ICOS+ |
| **61T** | Lymphocytes CD3+CD8+/ CD8+CD28+/ CD8+CD28+KLRG1+ |
| **62T** | Lymphocytes CD3+CD8+/ CD8+CD28+/ CD8+CD28+PD-1+ |
| **63T** | Lymphocytes CD3+CD8+/ CD8+CD28+/ Q1:CD57- KLRG1+ |
| **64T** | Lymphocytes CD3+CD8+/ CD8+CD28+/ Q2:CD57+ KLRG1+ |
| **65T** | Lymphocytes CD3+CD8+/ CD8+CD28+/ Q3:CD57+ KLRG1- |
| **66T** | Lymphocytes CD3+CD8+/ CD8+CD28+/ Q4:CD57- KLRG1- |
| **67T** | Lymphocytes CD3+CD8+/ CD8+CD28+/ Q5:CD57- PD-1+ |
| **68T** | Lymphocytes CD3+CD8+/ CD8+CD28+/ Q6:CD57+ PD-1+ |
| **69T** | Lymphocytes CD3+CD8+/ CD8+CD28+/ Q7:CD57+ PD-1- |
| **70T** | Lymphocytes CD3+CD8+/ CD8+CD28+/ Q8:CD57- PD-1- |
| **71T** | Lymphocytes CD3+CD8+/ CD8+CD57+ |
| **72T** | Lymphocytes CD3+CD8+/ CD8+ICOS+ |
| **73T** | Lymphocytes CD3+CD8+/ CD8+KLRG1+ |
| **74T** | Lymphocytes CD3+CD8+/ CD8+NAÏVE |
| **75T** | Lymphocytes CD3+CD8+/ CD8+PD-1+ |
| **76T** | Lymphocytes CD3+CD8+/ CD8+TIGIT+ |
| **77T** | Lymphocytes CD3+CD8+/ CD28-CD8+ |
| **78T** | Lymphocytes CD3+CD8+/ CD28-CD8+/ CD8+CD28-CD57+ |
| **79T** | Lymphocytes CD3+CD8+/ CD28-CD8+/ CD8+CD28-ICOS+ |
| **80T** | Lymphocytes CD3+CD8+/ CD28-CD8+/ CD8+CD28-KLRG1+ |
| **81T** | Lymphocytes CD3+CD8+/ CD28-CD8+/ CD8+CD28-PD-1+ |
| **82T** | Lymphocytes CD3+CD8+/ CD28-CD8+/ CD8+CD28-TIGIT+ |
| **83T** | Lymphocytes CD3+CD8+/ CD28-CD8+/ Q1: CD57- KLRG1+ |
| **84T** | Lymphocytes CD3+CD8+/ CD28-CD8+/ Q2: CD57+ KLRG1+ |
| **85T** | Lymphocytes CD3+CD8+/ CD28-CD8+/ Q3: CD57+ KLRG1- |
| **86T** | Lymphocytes CD3+CD8+/ CD28-CD8+/ Q4: CD57- KLRG1- |
| **87T** | Lymphocytes CD3+CD8+/ CM CD8+ |
| **88T** | Lymphocytes CD3+CD8+/ EFF CD8+ |
| **89T** | Lymphocytes CD3+CD8+/ EFF CD8+/ CD8+ EFF CD28- |
| **90T** | Lymphocytes CD3+CD8+/ EFF CD8+/ CD8+ EFF CD28-/ CD8+ EFF CD28-CD57+ |
| **91T** | Lymphocytes CD3+CD8+/ EFF CD8+/ CD8+ EFF CD28-/ CD8+ EFF CD28-KLRG1+ |
| **92T** | Lymphocytes CD3+CD8+/ EFF CD8+/ CD8+ EFF CD28-/ CD8+ EFF CD28-PD-1+ |

| | |
|---|---|
| **93T** | Lymphocytes CD3+CD8+/ EFF CD8+/ CD8+ EFF CD28-/ CD8+ EFF CD28-TIGIT+ |
| **94T** | Lymphocytes CD3+CD8+/ EFF CD8+/ CD8+ EFF CD28-/ Q2: CD57+ KLRG1+ |
| **95T** | Lymphocytes CD3+CD8+/ EFF CD8+/ CD8+ EFF CD28-/ Q3: CD57+ KLRG1- |
| **96T** | Lymphocytes CD3+CD8+/ EFF CD8+/ CD8+ EFF CD28-/ Q4: CD57- KLRG1- |
| **97T** | Lymphocytes CD3+CD8+/ EFF CD8+/ CD8+ EFF CD28-/ Q5: CD57- PD-1+ |
| **98T** | Lymphocytes CD3+CD8+/ EFF CD8+/ CD8+ EFF CD28-/ Q6: CD57+ PD-1+ |
| **99T** | Lymphocytes CD3+CD8+/ EFF CD8+/ CD8+ EFF CD28-/ Q7: CD57+ PD-1- |
| **100T** | Lymphocytes CD3+CD8+/ EFF CD8+/ CD8+ EFF CD28-/ Q8: CD57- PD-1- |
| **101T** | Lymphocytes CD3+CD8+/ EFF CD8+/ CD8+ EFF CD28-/ Q9: CD57- ICOS+ |
| **102T** | Lymphocytes CD3+CD8+/ EFF CD8+/ CD8+ EFF CD28-/ Q10: CD57+ ICOS+ |
| **103T** | Lymphocytes CD3+CD8+/ EFF CD8+/ CD8+ EFF CD28-/ Q11: CD57+ ICOS- |
| **104T** | Lymphocytes CD3+CD8+/ EFF CD8+/ CD8+ EFF CD28-/ Q12: CD57- ICOS- |
| **105T** | Lymphocytes CD3+CD8+/ EFF CD8+/ CD8+ EFF CD28-/ Q1: CD57- KLRG1+ |
| **106T** | Lymphocytes CD3+CD8+/ EFF CD8+/ CD8+ EFF CD57+ |
| **107T** | Lymphocytes CD3+CD8+/ EFF CD8+/ CD8+ EFF PD-1+ |
| **108T** | Lymphocytes CD3+CD8+/ EFF CD8+/ CD8+ EFF TIGIT+ |
| **109T** | Lymphocytes CD3+CD8+/ EFF CD8+/ Q1: CD57- TIGIT+ |
| **110T** | Lymphocytes CD3+CD8+/ EFF CD8+/ Q2: CD57+ TIGIT+ |
| **111T** | Lymphocytes CD3+CD8+/ EFF CD8+/ Q3: CD57+ TIGIT- |
| **112T** | Lymphocytes CD3+CD8+/ EFF CD8+/ Q4: CD57- TIGIT- |
| **113T** | Lymphocytes CD3+CD8+/ EM CD8+ |
| **114T** | Lymphocytes CD3+CD8+/ EM CD8+/ CD8+ EM CD57+ |
| **115T** | Lymphocytes CD3+CD8+/ EM CD8+/ CD8+ EM ICOS+ |
| **116T** | Lymphocytes CD3+CD8+/ EM CD8+/ CD8+ EM KLRG1+ |
| **117T** | Lymphocytes CD3+CD8+/ EM CD8+/ CD8+ EM PD-1+ |
| **118T** | Lymphocytes CD3+CD8+/ EM CD8+/ CD8+ EM TIGIT+ |
| **119T** | Lymphocytes CD3+CD8+/ EM CD8+/ CD28- CD45RO+ |
| **120T** | Lymphocytes CD3+CD8+/ EM CD8+/ CD28- CD45RO+/ CD8+ EM CD28-CD57+ |
| **121T** | Lymphocytes CD3+CD8+/ EM CD8+/ CD28- CD45RO+/ CD8+ EM CD28-KLRG1+ |
| **122T** | Lymphocytes CD3+CD8+/ EM CD8+/ CD28- CD45RO+/ CD8+ EM CD28-PD-1+ |
| **123T** | Lymphocytes CD3+CD8+/ EM CD8+/ CD28- CD45RO+/ Q2: CD57+ KLRG1+ |
| **124T** | Lymphocytes CD3+CD8+/ EM CD8+/ CD28- CD45RO+/ Q3: CD57+ KLRG1- |
| **125T** | Lymphocytes CD3+CD8+/ EM CD8+/ CD28- CD45RO+/ Q4: CD57- KLRG1- |
| **126T** | Lymphocytes CD3+CD8+/ EM CD8+/ CD28- CD45RO+/ Q5: CD57- PD-1+ |
| **127T** | Lymphocytes CD3+CD8+/ EM CD8+/ CD28- CD45RO+/ Q6: CD57+ PD-1+ |
| **128T** | Lymphocytes CD3+CD8+/ EM CD8+/ CD28- CD45RO+/ Q8: CD57- PD-1- |
| **129T** | Lymphocytes CD3+CD8+/ EM CD8+/ CD28- CD45RO+/ Q9: ICOS- TIGIT+ |
| **130T** | Lymphocytes CD3+CD8+/ EM CD8+/ CD28- CD45RO+/ Q10: ICOS+ TIGIT+ |
| **131T** | Lymphocytes CD3+CD8+/ EM CD8+/ CD28- CD45RO+/ Q11: ICOS+ TIGIT- |
| **132T** | Lymphocytes CD3+CD8+/ EM CD8+/ CD28- CD45RO+/ Q12: ICOS- TIGIT- |
| **133T** | Lymphocytes CD3+CD8+/ EM CD8+/ CD28- CD45RO+/ Q13: CD57- TIGIT+ |
| **134T** | Lymphocytes CD3+CD8+/ EM CD8+/ CD28- CD45RO+/ Q14: CD57+ TIGIT+ |
| **135T** | Lymphocytes CD3+CD8+/ EM CD8+/ CD28- CD45RO+/ Q15: CD57+ TIGIT- |
| **136T** | Lymphocytes CD3+CD8+/ EM CD8+/ CD28- CD45RO+/ Q16: CD57- KLRG1- |
| **137T** | Lymphocytes CD3+CD8+/ EM CD8+/ CD28- CD45RO+/ Q1: CD57- KLRG1+ |
| **138T** | Lymphocytes CD3+CD8+/ EM CD8+/ Q1: CD57- PD-1+ |
| **139T** | Lymphocytes CD3+CD8+/ EM CD8+/ Q2: CD57+ PD-1+ |
| **140T** | Lymphocytes CD3+CD8+/ EM CD8+/ Q3: CD57+ PD-1- |
| **141T** | Lymphocytes CD3+CD8+/ EM CD8+/ Q4: CD57- PD-1- |
| **HRV** | Horvath’s clock |
| **TL** | Telomere Length’s clock |
| **BLU** | BLUP clock |
| **SHM** | Shannon’s Entropy |

Summary representing the code for each biomarker used in the network analysis and the corresponding mediator.
