## Supplementary Figures for "Immunosenescence profile is associated with increased susceptibility to severe COVID-19"

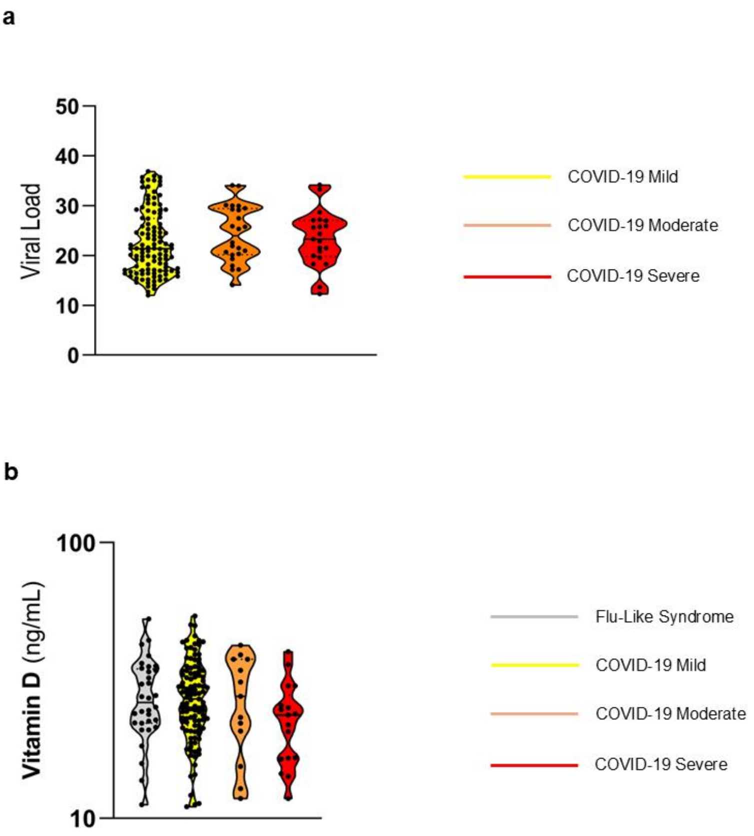


**Suppl. Figure 1 – The viral load and the vitamin D do not differ between levels of severity of COVID-19.** Differences in viral load and serological vitamin D in individuals with different clinical forms of COVID-19. The cohort used for this analysis was the composed by individuals from Belo Horizonte, São Paulo e Governador Valadares. (A) Viral load analysis included individual with mild COVID-19 presenting different clinical forms: mild (n = 27), moderate (n = 6) and severe (n = 8). (B) Vitamin D levels of COVID-19 patients with different clinical forms were compared within groups and with Flu-like syndrome patients (n = 34). The clinical forms considered were: mild (n = 111), moderate (n = 14), and severe (n = 25). Samples were previously normalized, and outliers were excluded using the ROUT test. Mann-Whitney test was performed individually for each group. Lines were used to represent which groups were compared and asterisks to represent statistical significance ($p \leq 0.05$, $^{**}p \leq 0.01$, $^{***}p \leq 0.001$ or $^{****}p < 0.0001$). All groups presented in this figure were matched by sex as well as age and they were composed of individuals at the initial stage of infection (1-4 days of symptoms).

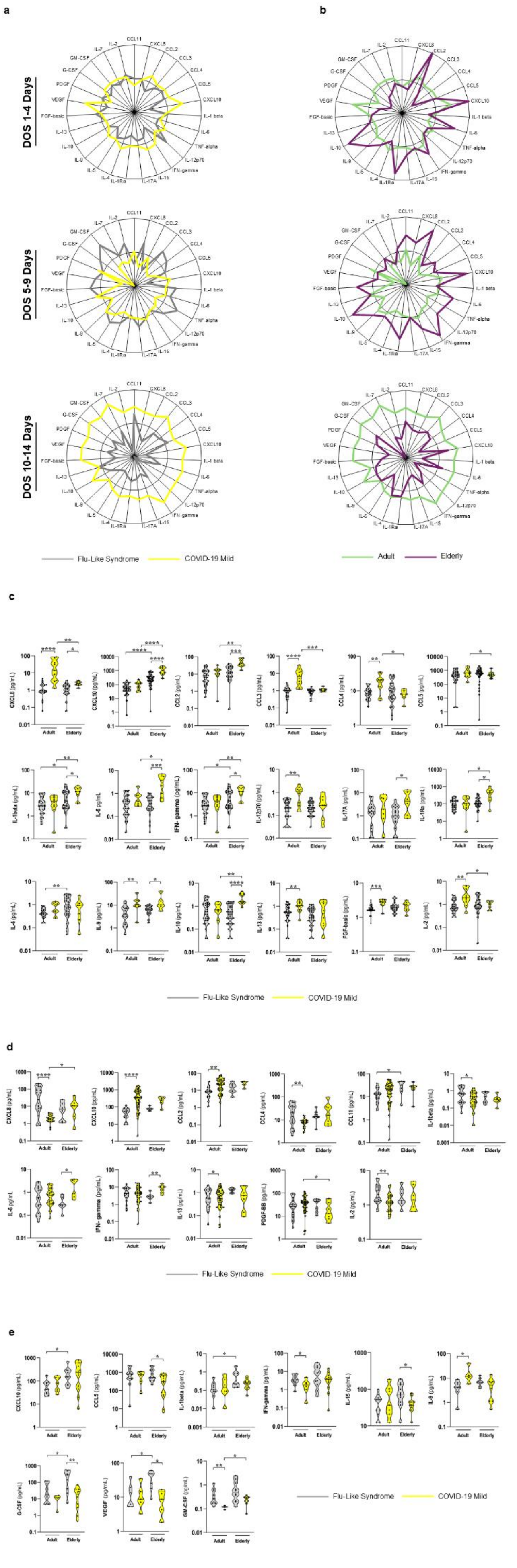

a
b
DOS 1-4 Days
DOS 5-9 Days
DOS 10-14 Days
Flu-Like Syndrome
COVID-19 Mild
Adult
Elderly
c
Flu-Like Syndrome
COVID-19 Mild
d
Flu-Like Syndrome
COVID-19 Mild
e
Flu-Like Syndrome
COVID-19 Mild

**Suppl. Figure 2 – Individual with mild COVID-19 presents a higher immune activation between 10 to 14 days of symptoms when compared to FLS patients, while the elderly present higher global inflammatory profile during early stages of the disease when compared to adults. (A)** Radar chart with the frequency of high mediator producers among individuals from Belo Horizonte and São Paulo according to the days since symptom onset. 1 to 4 days with Flu-like syndrome (FLS) (n = 50) and mild-COVID-19 (n = 41); 5 to 9 days with Flu-like syndrome (FLS) (n = 37) and mild-COVID-19 (n = 53); 10 to 14 days with Flu-like syndrome (FLS) (n = 14) and mild-COVID-19 (n = 15). All groups presented in this figure were matched by sex as well as age. (B) Radar chart with the frequency of high mediator producers among individuals from Belo Horizonte and São Paulo according to the age range and days since symptom onset: adults (n = 31) and elderly (n = 6) from 1 to 4 days of symptoms; adults (n = 46) and elderly (n = 7) from 5 to 9 days of symptoms; and adults (n = 7) and elderly (n = 7) from 10 to 14 days of symptoms. Differences in plasma mediator concentrations in adults and elderly with FLS and mild COVID-19 in different time points of symptom onset: (C) 1 to 4 days (n = 103); (D) 5 to 9 days (n = 91); (E) 10 to 14 days (n = 30). Samples were previously normalized, and outliers were excluded using the ROUT test. Mann-Whitney test was performed individually for each group. Lines were used to represent which groups were compared and asterisks to represent statistical significance ($p \leq 0.05$, $^{**}p \leq 0.01$, $^{***}p \leq 0.001$ or $^{****}p < 0.0001$).

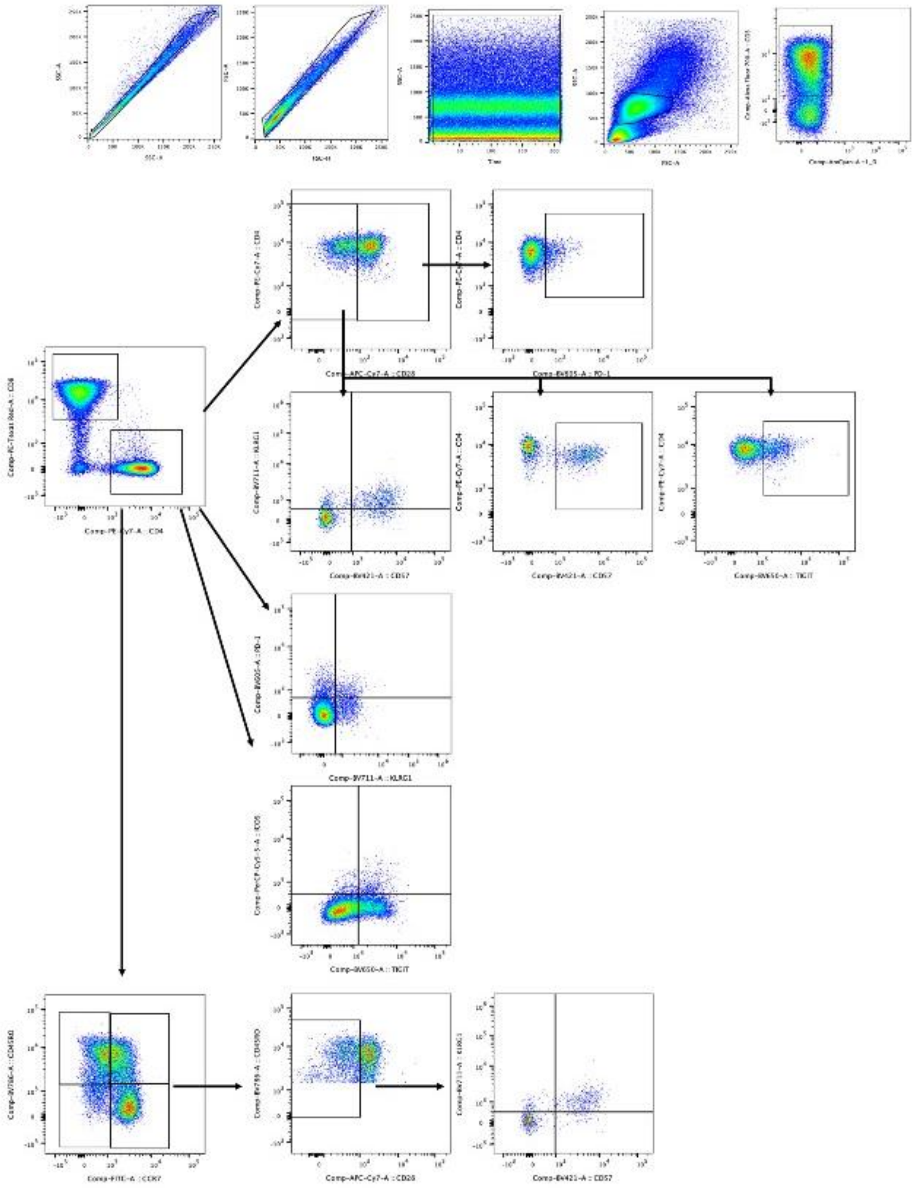

**Suppl. Figure 3 – Gating strategy for the identification of the indicated T cell subsets in peripheral blood mononuclear cells (PBMCs).**

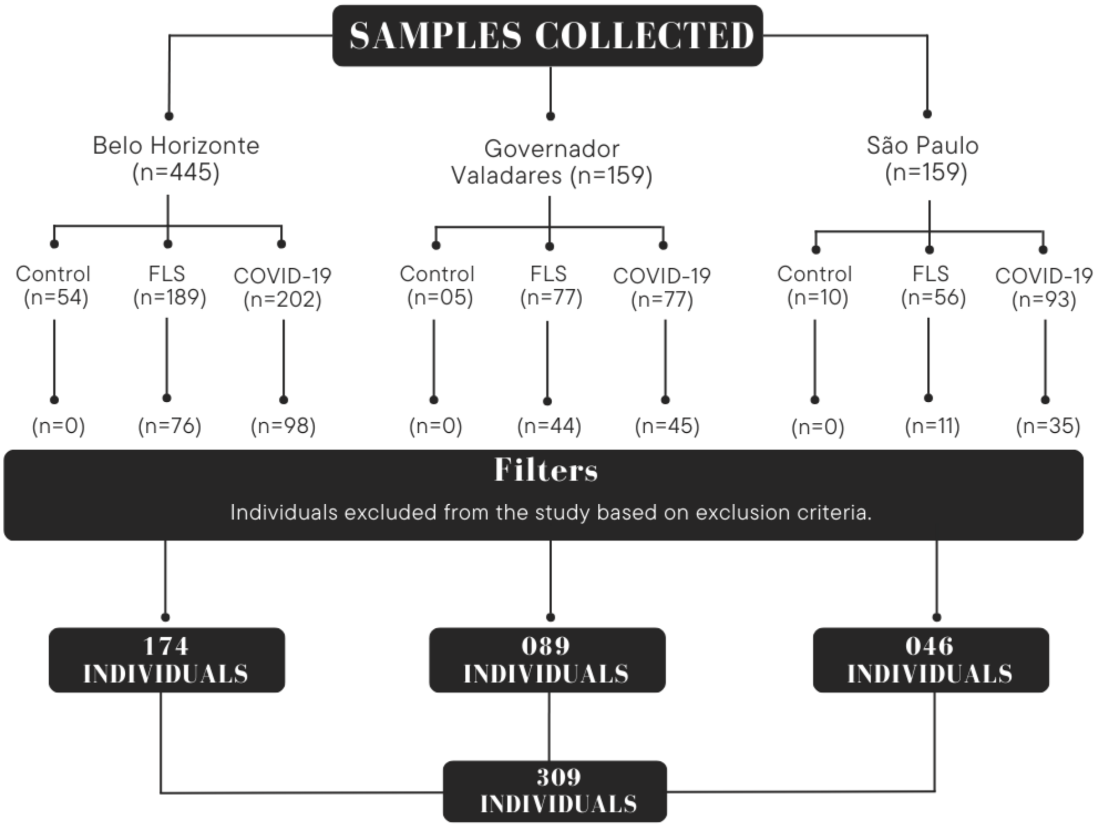


**Suppl. Figure 4 – Sample selection flowchart (Study Design)**
